# Pathological slow-wave activity and impaired working memory binding in post-traumatic amnesia

**DOI:** 10.1101/2021.12.24.21268337

**Authors:** Emma-Jane Mallas, Nikos Gorgoraptis, Sophie Dautricourt, Yoni Pertzov, Gregory Scott, David J. Sharp

**Affiliations:** Department of Brain Sciences, Imperial College London, London, UK; UK Dementia Research Institute, Care Research and Technology Centre, Imperial College London, London, UK; Inserm UMR-S U1237, Caen-Normandie University, GIP Cyceron, Caen, France; Visual Cognition Lab, The Hebrew University of Jerusalem, Jerusalem, Israel; Royal British Legion Centre for Blast Injury Studies, Department of Bioengineering, Imperial College London, London, UK

**Author notes:** These authors contributed equally to this work (Gregory Scott, David J. Sharp). ***Correspondence to:*** Dr Gregory Scott, UKDRI Care Research and Technology Centre, Sir Michael Uren Hub, Imperial College London, 86 Wood Lane, London W12 0BZ.

**Keywords:** post-traumatic amnesia, traumatic brain injury, working memory, binding, EEG

## Abstract

The mechanism by which information is bound together in working memory is a central question for cognitive neuroscience. This binding is transiently disrupted during periods of post-traumatic amnesia following significant head injuries. The reason for this impairment is unclear but may be due to electrophysiological changes produced by head impacts. These are common and include pathological low frequency activity, which is associated with poorer neurological outcomes and may disrupt cortical communication. Here, we investigate associative memory binding during post-traumatic amnesia and test the hypothesis that misbinding is caused by a disruption in cortical communication produced by the pathological slowing of brain activity. Thirty acute moderate-severe traumatic brain injury patients (mean time since injury = 10 days) and 26 healthy controls were tested with a precision working memory paradigm that required the association of object and location information. A novel entropy ratio measure was calculated from behavioural performance. This provided a continuous measure of the degree of misbinding and the influence of distracting information. Resting state EEG was used to assess the electrophysiological effects of traumatic brain injury. Patients in post-traumatic amnesia showed abnormalities in working memory function and made significantly more misbinding errors than patients who were not in post-traumatic amnesia and controls. Patients showed a higher entropy ratio in the distribution of spatial responses, indicating that working memory recall was abnormally biased by the locations of non-target items suggesting a specific impairment of object and location binding. Slow wave activity was increased following traumatic brain injury. Increases in the delta-alpha ratio indicative of an increase in low frequency power specifically correlated with binding impairment in working memory. In contrast, although connectivity was increased in the theta band and decreased in the alpha band after traumatic brain injury, this did not correlate with working memory impairment. Working memory and electrophysiological abnormalities both normalised at six-month follow-up, in keeping with a transient increase in slow-wave activity causing post-traumatic amnesia that impaired working memory function. These results show that patients in post-traumatic amnesia show high rates of working memory misbinding that are associated with a pathological shift towards lower frequency oscillations.

## Introduction

Post-traumatic amnesia (PTA) is a transient state of cognitive impairment following traumatic brain injury (TBI) characterised by disorientation, confusion, and the inability to encode new memories. A period of PTA is usually short lived, but can vary from seconds to months and may persist throughout inpatient care.^1,2^ PTA duration is a marker of TBI severity with longer periods associated with poorer functional outcomes.^3,4^ We have previously shown that working memory impairments during PTA are associated with a disruption of functional connectivity across the limbic system, measured by functional MRI.^5^ Reductions in connectivity between the parahippocampal gyrus and the posterior cingulate cortex were associated with working memory impairments, which resolved when patients emerged from PTA. This suggests that working memory impairments seen during PTA result from a disruption to cortical communication between brain regions involved in encoding the information held in working memory.

Post-traumatic amnesia can lead to a range of working memory impairments.^5–7^ In the case of visual working memory, a range of information about an object such as its identity and location can be encoded and bound together, with binding a particularly sensitive measure of working memory impairment in various disease states.^8–12^ Classic span tasks such as the N-back are sensitive to general impairments of working memory, but don’t allow the binding of information to be specifically studied. Recently, precision working memory tasks have been developed that allow recognition memory for object identity and spatial location to be measured separately from the binding of this information e.g. providing separate tests of whether an object was remembered, and whether the spatial location of that object was remembered.^9^ This allows detailed investigation of the type of working memory impairment seen in PTA, which we use to investigate whether the failure to bind information in working memory is associated with electrophysiological abnormalities that potentially disrupt cortical communication.

The neural basis of working memory impairments can be studied using EEG. Neural oscillations are considered central to working memory function.^13^ For example, synchronising the phase of neural oscillations across distinct brain regions supports information exchange between prefrontal and temporal regions during working memory.^14–16^ Additionally, the cross-frequency coupling of phase and amplitude (PAC) is key to hippocampal memory function,^17–20^ and the strength of theta-gamma PAC is related to working memory load, successful memory encoding, working memory maintenance and associative binding.^21–26^ The precise synchronisation of neural oscillations necessary for working memory function may be sensitive to disruption by large electrophysiological abnormalities that are often seen transiently after head injury.

Increased low-frequency oscillations are seen in many disease states, including TBI.^27–31^ These can be quantified by measuring power within EEG frequency bands. Relative changes are often described using the delta-alpha ratio (DAR), which reflects the difference in contribution between delta (0-4 Hz) and alpha (8-13Hz). The DAR correlates with clinical outcomes and cognitive function following acquired brain injury.^32–34^ Increases in DAR are associated with poorer neurological outcome including personality change, depressive symptoms and cognitive impairment following head injury.^30,31,35–40^ There is also evidence that reducing DAR using transcranial electrical stimulation can improve cognitive performance after TBI.^41^ Synchronisation disturbances are also present following TBI across a range of frequency bands including decreased gamma connectivity.^42–47^ Reduced connectivity across theta, alpha and beta (13-30 Hz) bands has been associated with working memory impairment after TBI^48,49^ and improvements in cognition associated with reduced delta connectivity.^47^ PAC is abnormal following TBI suggesting that cross-frequency coupling is also disrupted.^50^ Taken together, this suggests that a shift to pathological low-frequency power after TBI may be mechanistically important for working memory disturbance by disrupting the neural oscillations than support the maintenance of information in working memory.

Here for the first time, we investigate working memory binding impairments in an acute cohort of moderate-severe TBI patients with and without PTA. A precision recall visual working memory paradigm is used to sensitively quantify the degree of misbinding of object and location information within working memory. A novel entropy ratio measure was calculated from behavioural performance, providing a continuous measure of the degree of misbinding and the influence of distracting spatial information. Electrophysiological abnormalities seen following head injuries are quantified using EEG. We test the following hypotheses: 1) patients in PTA will show impairments of working memory binding, demonstrated by misbinding errors and an increased entropy ratio of spatial response distribution; 2) PTA will be associated with increased low frequency oscillations, compared to controls and TBI patients not in PTA; 3) working memory binding impairment will correlate with increased low frequency oscillations reflected in increased DAR; 4) PTA will be associated with abnormal long-range theta phase synchronisation and theta-gamma PAC between frontal and parietal-temporal channels; 5) Emergence from PTA will be associated with a reduction in working memory binding errors and a normalisation of EEG measures.

## Materials and methods

### Participant demographics and clinical details

#### Traumatic brain injury group

Thirty patients (25 males, five females, mean age 40.73, range 17-73 years) admitted with a recent history of TBI were recruited from the Major Trauma Ward, St Mary’s Hospital, London, UK. All patients had moderate-severe injuries according to the Mayo Classification system for TBI severity (Supplementary Table 1).^51^ Injuries were secondary to road traffic accidents (40.0%), falls (33.3%), assault (20.0%) and sports injury (6.67%). Patients were included if they were between the ages of 16-80 and clinically stable. Exclusion criteria were as follows: premorbid psychiatric or neurological illness; history of other significant TBI; current or previous drug or alcohol abuse; pregnancy or breastfeeding; significant language or visuospatial impairments. For the EEG part of the study, neurosurgical intervention (or other contraindication to scalp EEG) was also an exclusion criterion.

Written informed consent was obtained for patients judged to have capacity according to the Declaration of Helsinki. Patients in PTA who were judged not to have capacity were deemed unable to give informed consent for participation in the study. In this case, written assent was obtained as well as informed written assent by a caregiver on the patient’s behalf. Informed consent was gained retrospectively once patients emerged from PTA. No patients withdrew consent. The study was approved by the West London Research Ethics Committee (09/HO707/82).

#### Control group

26 healthy controls (20 males, 6 females, mean age 28.96, range 18-70 years) were recruited. Participants had no history of psychiatric or neurological illness, previous TBI or alcohol or substance misuse. All participants gave written informed consent.

### Protocol

At baseline, all participants underwent neuropsychological assessment, precision working memory task and resting state EEG. Additionally, patients underwent PTA assessment according to the Westmead Post Traumatic Amnesia Scale (WPTAS).^52^ The WPTAS is a 12-item scale with seven items that assess orientation and five items that assess memory. Patients must score full marks (12/12) for three consecutive days before they are deemed to no longer be in PTA. PTA duration is calculated as the time between injury and the first of these successful consecutive assessments. Patients were on average within 10 days of injury (range 1-32 days) and were divided into two groups (PTA+ and PTA-). Patients were invited to attend follow-up assessment within six months of hospital discharge, at which the baseline protocol was repeated. Controls were assessed at one timepoint.

Where possible, participants took part in all aspects of the study. However, due to the nature of recruitment within an acute clinical setting this was not always possible or appropriate. A subsample of 17 patients and 21 controls underwent resting state EEG. Of these, there was one patient and one control who did not complete the precision working memory task.

#### Neuropsychological assessment

All participants completed a detailed neuropsychology assessment with a focus on episodic and working memory. Immediate and delayed verbal recall were assessed using The Logical Memory I and II subtests of the Wechsler Memory Scale, third edition.^53^ Immediate and delayed visuospatial memory were assessed using the Brief Visuospatial Memory Test-Revised.^54^ A battery of computerised tests based on classical paradigms from the cognitive psychology literature was delivered on a tablet device using a custom-programmed application. Details of each task have been previously reported.^55^ In brief: Visuospatial Working Memory, based on a task from the non-human primate literature was used to assess visuospatial working memory^56^; Paired Associates, based on a paradigm commonly used to assess memory impairments in aging clinical populations was used to assess object-location association memory^57^; Spatial span, based on the Corsi Block Tapping Task was used to assess spatial short term memory capacity^58^; Self-Ordered Search was used to measure strategy during search behaviour, based on an existing task^59^; Feature Match, based on the classical feature search task was used to measure attentional processing.^60^

##### Statistical analysis of neuropsychological data

One-way ANOVAs were used to identify group effects at baseline. Post-hoc independent samples t-tests, using false discovery rate (FDR) multiple comparisons corrections were performed to determine which pairwise comparisons were driving any significant main effects. Linear mixed-effects models were used to assess longitudinal changes between baseline and follow-up in which group and time point were defined as fixed effects and subject was defined as a random effect. Post-hoc paired samples t-tests were used to investigate any significant main effects or interactions. Unless otherwise stated all statistical analysis was performed using R (Version 1.3.1056).

#### Experimental task paradigm

Participants completed a precision recall working memory task (Figure 1) based on Pertzov et al.^9^ The stimuli consisted of 60 fractal images.^61^ Fractals were used as complex visual objects that cannot be readily verbalised. A maximum width and height of 120 pixels was used. Stimuli were presented on an interactive touch-sensitive screen with a 1920 × 1080 pixel matrix (Dell Corp. Ltd). The experiment consisted of 80 trials (20 1 item; 60 2 item). Object location was determined by a Matlab script (MathWorks) in a random manner at any possible location on the screen with exceptions: objects were never located within 600 pixels of each other within a single trial. They were positioned within a minimum distance of 200 pixels from the screen edge. The threshold for the distance at which the response matched the target (or distractor) was set to 200 pixels.

**Figure 1.**
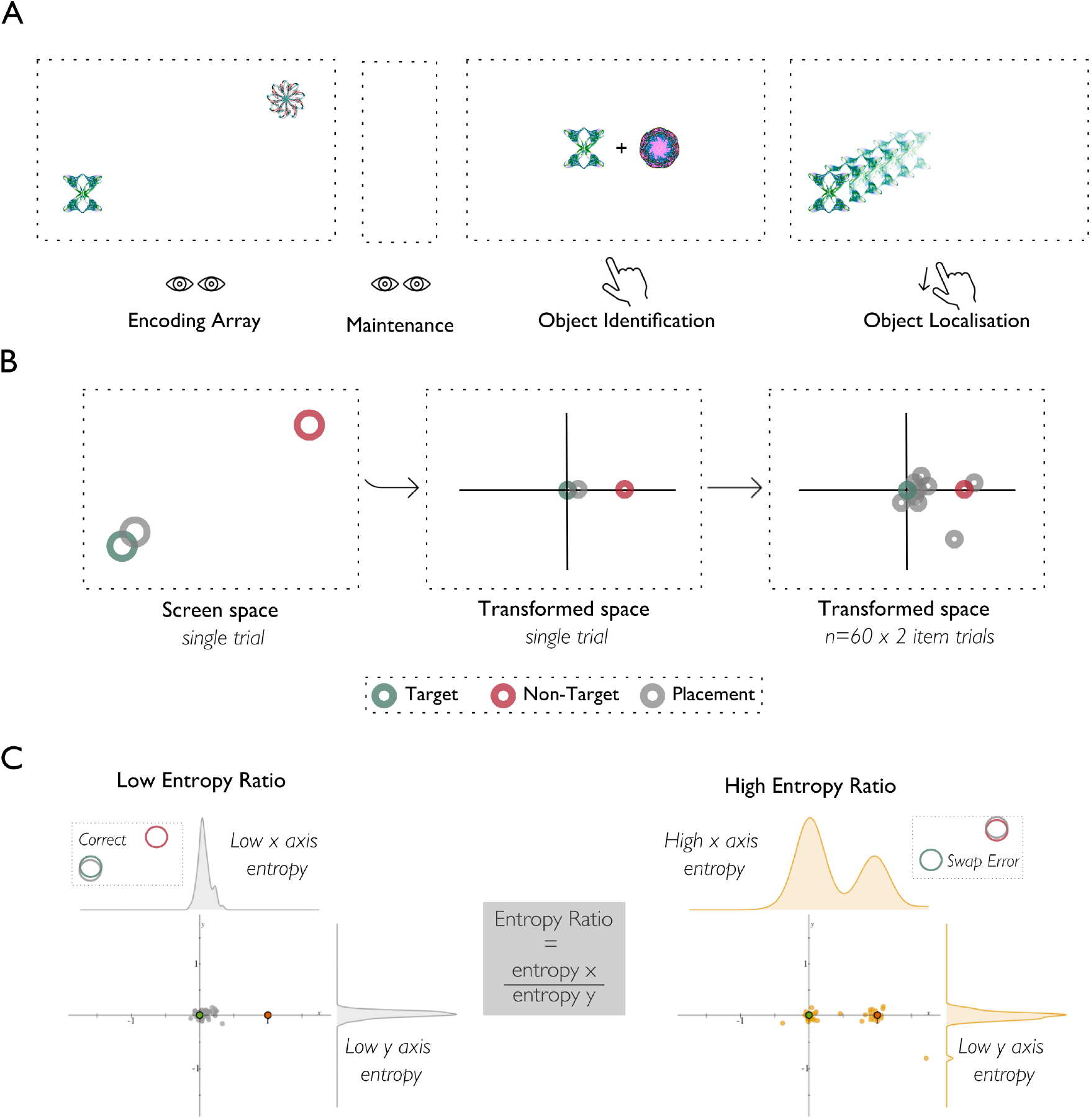
Precision working memory task. **(A)** Experimental task paradigm. One or two fractal objects are shown (‘Encoding Array’) prior to a delay of 2 seconds (‘Maintenance’). One of the original objects (Target) is then displayed alongside a novel object (Non-Target), which had not appeared in the encoding array. Participants are required to select the object they recall (‘Object Identification’) and move it to its remembered location (‘Object Localisation’). **(B)** Transformation of data from screen space to transformed space in which the target lies at the origin, the non-target at 1,0 (x, y) and the object placement for each trial is plotted in relation to these constants. **(C)** The placements across all trials are quantified using a ratio of the entropy of the distribution of responses across the x and y axes. A low entropy ratio indicates responses are clustered around the target indicating correct responses and a high entropy ratio indicates a second cluster around the non-target indicating misbinding errors. For random responses (not shown), entropy would increase across both x and y axes.

##### Task procedure

Each trial began with a central fixation point followed by the memory array consisting of 1 or 2 distinct fractals presented for 2 seconds. A blank screen was displayed for 2 seconds (maintenance period) after which the object identification stage began in which two fractals were displayed alongside one another in the centre of the screen. One fractal had been present in the memory array of that trial, the other was a foil item that was not present in the memory array. The foil item was part of the task stimuli and therefore was not unfamiliar but appeared in other trials throughout the experiment. Participants were required to touch the item that they remembered from the memory array and then drag it to the remembered location. Localisation performance was only analysed in trials in which the object identity was correctly remembered.

##### Task analysis

Data from the precision working memory task were analysed using (a) a thresholded approach in which a correct localisation was considered to be within 200 pixels of the target (or a misbinding error within 200 pixels of the non-target) to calculate a proportion of misbinding errors and (b) a novel, threshold-free approach to study the distribution of responses across a transformed space, defined by the relative locations of target versus non-target items (Figure 1B). In this transformed space the target position is located at the origin and the non-target at 1,0 (*x,y*). Therefore, responses deviating from the origin along the x-axis indicate a spatial bias by the non-target item location, as happens when the target identity is misbound with the non-target location. In contrast, responses deviating along the y-axis do not indicate a spatial bias by the non-target location but are in-keeping with imprecision in remembering the correct location, or with random error. The responses in this transformed space are binned across ten uniformly spaced bins. A measure of entropy of the distribution of responses across each axis is then obtained, and a ratio of x-axis over y-axis entropy is calculated. This entropy ratio serves as a quantitative measure of response bias by the location of non-target items, and the direction of the effect (bias toward vs. bias away from the non-target item) becomes apparent by plotting the responses in the transformed space (Figure 1C). This approach allows the influence of the non-target on the remembered location to be precisely examined without dichotomising responses based on arbitrarily defined thresholds. The number of bins chosen did not impact the difference in entropy ratio between groups (Supplementary Fig.1). The design of this paradigm thus allowed us to measure the distinct processes of object-location associative memory independently to quantify the types of error made, i.e., failure to remember the location or failure to bind.

#### EEG data acquisition

EEG data were acquired using a 32-channel active electrode standard actiCAP positioned with Cz at the midline with electrodes grounded to FPz and referenced to Fz. Signals were amplified using the actiCHamp system and data recorded using the BrainVision Recorder software (Brain Products) at a sampling rate of 1000 Hz.

#### EEG pre-processing

Raw EEG data were exported from BrainVision into Matlab via EEGLAB. Data were pre-processed, cleaned and quality assessed using the Harvard Automated Processing Pipeline for EEG (HAPPE), an automated pre-processing pipeline specifically developed for high artefact data as would be expected in acute TBI patients. HAPPE has been shown to be superior in optimising signal to noise ratio compared to manual editing in clinical data.^62^ The following pre-processing steps were taken using the HAPPE pipeline: Data were high-pass filtered at 1Hz, low-pass filtered at 100Hz and band-pass filtered at 1-249Hz as recommended for HAPPE processing. Electrical line noise at 50Hz was removed using the CleanLine multi-taper approach.^63^ Bad channels (i.e. those with poor signal quality) were rejected using joint probability evaluation and those exceeding 3 SD from the mean were excluded from further analyses (to be later interpolated). Wavelet-enhanced independent component analysis (wICA) was performed to correct for EEG artefact while retaining the whole dataset to improve ICA decomposition. ICA was performed on the corrected data and components were rejected using the multiple artifact rejection algorithm (MARA).^64^ HAPPE rejects components with artifact probabilities > 0.5. Data were segmented into 5-second segments and subjected to amplitude-based (+/-40μV) and joint-probability (< 3 SDs relative to the activity of other segments) rejection to remove segments with remaining artefact. Previously rejected channels were repopulated using spherical interpolation. Data were re-referenced to average.

#### EEG analysis

Channels were grouped into four regions: frontal (‘*Fp1’, ‘Fp2’, ‘F3’, ‘F4’, ‘F7’, ‘F8’*), temporal *(‘T7’, ‘T8’, ‘TP9’, ‘TP10’, ‘FT9’, ‘FT10’)*, parietal (‘*Pz’, ‘P3’, ‘P4’, ‘P7’, ‘P8’*), occipital (‘*O1’, ‘O2’, ‘Oz’)*. Frequency bands were defined as follows: delta (0-4Hz); theta (4-8Hz); alpha (8-13Hz); beta (13-30Hz) and gamma (30-40Hz).

##### Normalised power

Power in each channel was calculated for each frequency band using multitaper frequency transformation normalised to total power across all five bands. Global power was calculated by averaging across all channels.

Statistical comparison of global normalised power was conducted using one-way independent measures ANOVA followed by post-hoc *t*-tests. All *p*-values were corrected using FDR method for multiple comparisons. Group-level statistical analysis of normalised power was also performed using a cluster-based permutation approach in the frequency/channel domain on the whole montage.^65^ Power was compared between groups at each channel using two-sided independent samples t-tests and results clustered according to spatial adjacency at *P*<0.05 using the maximum size criterion. Permutation distributions were generated using the Monte-Carlo method and 5000 random iterations, and corrected *p*-values were then obtained through comparison of observed data to the random distributions. Follow-up data was assessed as above. No cluster-based permutation analysis was performed on follow-up data.

##### Phase synchronisation

Phase synchronisation was quantified using the debiased weighed phase lag index (dwPLI) a measure of phase based functional connectivity. Phase lag index (PLI) calculates to what extent the phase of one signal is consistently lagging or leading relative to another signal, irrespective of the magnitude of the phase leads and lags.^66^ The weighted PLI is weighted by the imaginary component of the cross-spectrum to overcome issues with spuriously related connectivity which can arise due to volume conduction.^67^ A further debiasing term to correct for inflation due to small sample size was additionally implemented within the ft_connectivity_wpli.m function in FieldTrip.^68^ dwPLI is calculated across all frequencies within the band of interest for each channel pair.

To statistically compare connectivity at the group level we constructed 31×31 whole brain channel-wise connectivity matrices for each subject by averaging across dwPLI values for each channel pair across the frequency band. The network-based statistic (NBS) implemented in MATLAB was used to compare connectivity across groups using an independent samples *t*-test design.^69^ The NBS is a non-parametric statistical method in which values at every node are tested against the null-hypothesis and those surviving the primary threshold are entered for Monte-Carlo simulation permutation testing at every channel pair. The primary threshold was set to *z*=3.1 and 10,000 random permutations were conducted with a threshold of *P*<0.05.

##### Phase-amplitude coupling

To quantify the intensity of PAC between theta phase and gamma amplitude we computed the modulation index (MI).^70^ We estimated the phase of frequencies between 4-8 Hz (in steps of 1 Hz) in the frontal channel group and the amplitudes of frequencies between 30-40 Hz (in steps of 2 Hz) in parietal and temporal channel groups individually. The MI was calculated separately for each electrode within the respective channel groups.

To test for group differences in PAC, MI values were averaged across frontal-parietal and separately frontal-temporal channel groups for each participant and compared using independent samples *t*-tests. In patients who returned for follow-up, within-subjects *t*-tests were used to assess differences in the dwPLI and MI across time.

## Results

### Clinical demographics

Thirty moderate-severe TBI patients were recruited during their in-patient stay (age range 17-73 years; Table 1). Seventeen patients were in PTA at the time of enrolment (PTA+, mean WPTAS 9.18, *SD*=1.38). Thirteen patients were not in PTA (PTA-, WPTAS 12 for all). The PTA+ group had a longer PTA duration and longer hospital admission, but groups did not differ in time since injury (Table 1). PTA+ and PTA- groups were well matched for age (*P*=0.95) and gender (Table 1).

**Table 1.**
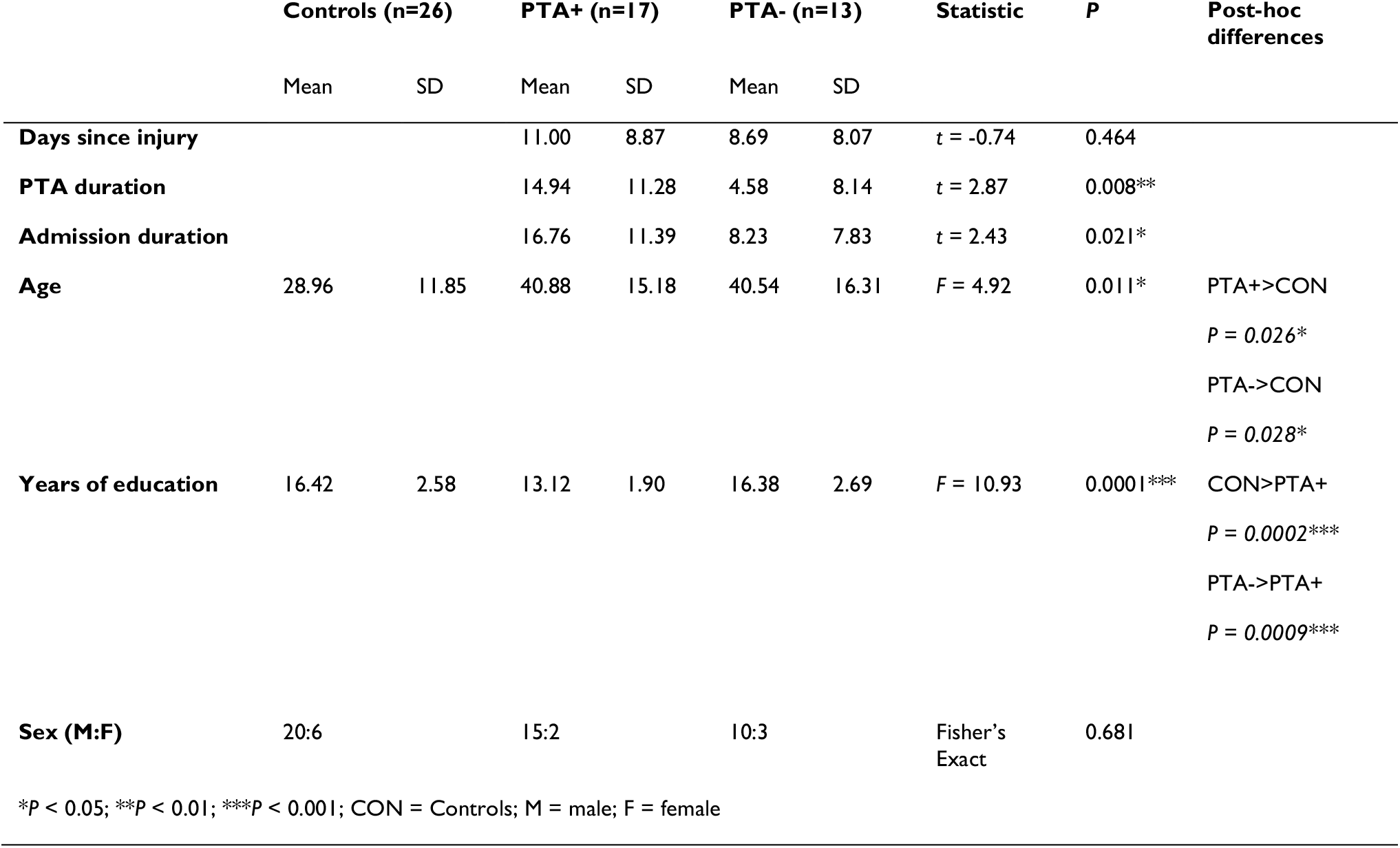
Clinical demographics for all participants.

|  | Controls (n=26) |  | PTA+ (n=17) |  | PTA- (n=13) |  | Statistic | P | Post-hoc differences |
| --- | --- | --- | --- | --- | --- | --- | --- | --- | --- |
|  | Mean | SD | Mean | SD | Mean | SD |  |  |  |
| **Days since injury** | | | 11.00 | 8.87 | 8.69 | 8.07 | $t = -0.74$ | 0.464 | |
| **PTA duration** | | | 14.94 | 11.28 | 4.58 | 8.14 | $t = 2.87$ | 0.008** | |
| **Admission duration** | | | 16.76 | 11.39 | 8.23 | 7.83 | $t = 2.43$ | 0.021* | |
| **Age** | 28.96 | 11.85 | 40.88 | 15.18 | 40.54 | 16.31 | $F = 4.92$ | 0.011* | PTA+>CON<br>$P = 0.026^*$<br>PTA->CON<br>$P = 0.028^*$ |
| **Years of education** | 16.42 | 2.58 | 13.12 | 1.90 | 16.38 | 2.69 | $F = 10.93$ | 0.0001*** | CON>PTA+<br>$P = 0.0002^{***}$<br>PTA->PTA+<br>$P = 0.0009^{***}$ |
| **Sex (M:F)** | 20:6 |  | 15:2 |  | 10:3 |  | Fisher's Exact | 0.681 |  |

\* $P < 0.05$ ; \*\* $P < 0.01$ ; \*\*\* $P < 0.001$ ; CON = Controls; M = male; F = female

Twenty-six control participants were recruited (age range 18–70 years; Table 1). ANOVA showed an effect of age across the groups driven by controls being slightly younger than both PTA+ and PTA- patients (Table 1). Additionally, there were group differences in years of education due to the PTA+ group having fewer years of education than controls and the PTA-group (Table 1).

### Neuropsychological performance

TBI patients generally had significant cognitive abnormalities (Supplementary Fig.1; Supplementary Table 2), with impairments in the following relative to controls: verbal memory (Immediate: *F*(2,34)=11.20, *P*<0.001; Delayed: *F*(2,34)=9.46, *P*<0.001), visuospatial memory (BVMT Immediate: *F*(2,37)=7.57, *P*=0.002; BVMT Delayed: *F*(2,37)=6.66, *P*=0.003; computerised: (*F*(2,45)=5.26, *P*=0.010), associative working memory (*F*(2,45)=10.09, *P*<0.001), spatial short term memory (*F*(2,44)=4.63, *P* =0.015), search strategy (*F*(2,45)=15.24, *P*<0.001) and attentional processing (*F*(2,45)=19.48, *P*<0.001). There were no group differences in retention rates between immediate and delayed recall for either verbal memory or visuospatial memory.

Patients in PTA also showed impairments compared to both PTA- patients and controls in the following: immediate verbal recall (PTA+ < PTA-: *t*(34)=2.66, *P*=0.018; PTA+ < CON: *t*(34)=-4.69, *P*<0.001), delayed visuospatial memory recall (PTA+ < PTA-: *t*(36)=2.26, *P*=0.045; PTA+ < CON: *t*(36)=-3.59, *P*=0.003) associative working memory (PTA+ < PTA-: *t*(45)=2.94, *P*=0.008; PTA+ < CON: *t*(45)=-4.43, *P*<0.001) and search strategy (PTA+ < PTA-: *t*(45)=4.11, *P*<0.001; PTA+ < CON: *t*(45)=-5.28, *P*<0.001). PTA patients also showed impairments compared to controls in delayed verbal memory recall (PTA+ < CON: (*t*(34) = -4.34,*P*<0.001), immediate visuospatial memory recall (PTA+ < CON: *t*(37) =-3.89, *P*=0.001), visuospatial working memory (PTA+ < CON: *t*(45)=-3.11,*P*=0.010), short term spatial memory (PTA+ <CON: *t*(44)=-3.04, *P*=0.012) and attentional processing (PTA+ < CON: *t*(45) = -6.10, *P*<0.001; PTA- < CON: *t*(45)=-3.72, *P*<0.001).

### Impaired working memory binding in post-traumatic amnesia

A precision working memory task was used to assess the binding of object and location information in working memory. All three groups identified the target with >90% accuracy, indicating that subjects understood the task regardless of whether they were in PTA (Figure 2A). As expected, accuracy decreased as working memory load increased, with two item trials less accurate than one item trials (*F*(1,51) = 55.32, *P*<0.001). There was also a group effect of object identification (*F*(2,51)=9.98, *P*<0.001): PTA+ patients showed more errors in identifying targets than PTA-patients (*t*(105)=3.49, *P*=0.001) and controls (*t*(105)=-5.46, *P*<0.001). A group by load (1 or 2 item trial) interaction was present of borderline significance (*F*(2,51)=2.89, *P*=0.065).

**Figure 2.**
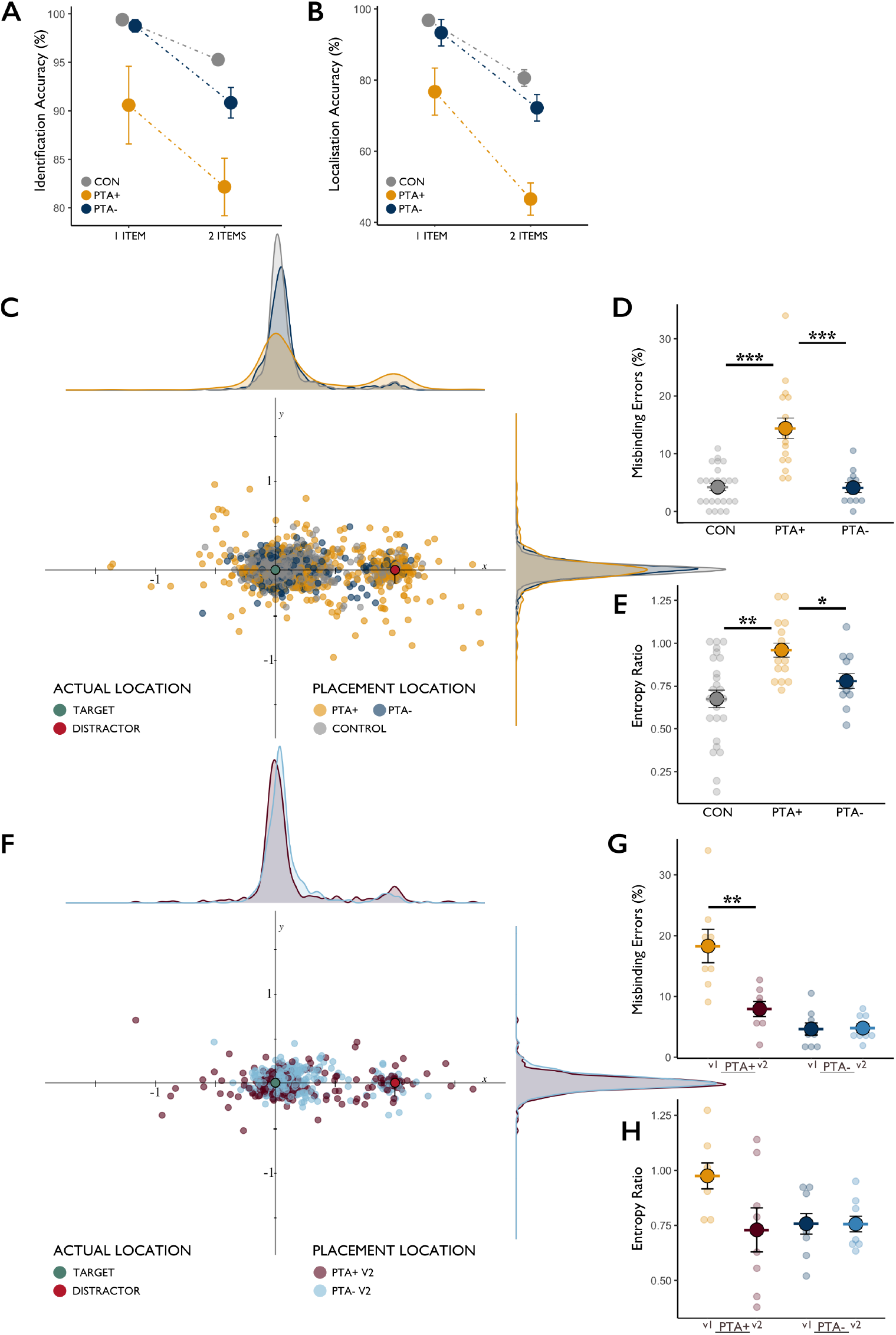
Performance on the object-location association precision working memory task in healthy controls and PTA+ and PTA-TBI patients at baseline and follow-up. **(A)** Object identification accuracy at baseline for 1 and 2 item trials. **(B)** Percentage of correctly identified trials that were moved to the correct location in 1 and 2 item trials at baseline. **(C)** Scatter-hist showing the distribution of location responses correctly identified trials in a normalised space in reference to the target and non-target locations at baseline. **(D)** Percentage of correctly identified trials that resulted in a misbinding error at baseline. **(E)** Entropy ratio of the x and y axes for the distribution of responses shown in panel C. **(F)** Scatter-hist showing the distribution of location responses for PTA+ and PTA-TBI patients that returned for follow-up. **(G)** Proportion of misbinding errors made in correctly identified trials by PTA+ and PTA-patients at baseline and follow-up. **(H)** Entropy ratio of distribution of responses as shown in panel F, in PTA+ and PTA-patients at baseline and follow-up. ***Significance at P < 0.001; ** significance at P < 0.01; * significance at P < 0.05. Error bars represent the standard error of the mean (SEM). CON = healthy controls.

Similar results were observed when considering the spatial accuracy of target placement (Figure 2B). A group by load interaction was present (*F*(2,51)=7.32, *P*=0.002), due to PTA+ patients showing less accurate placements compared to PTA-patients (*t*(105)=4.10, *P*<0.001) and controls (*t*(105) =-6.31,*P*<0.001).

Next, we examined the numbers of misbinding errors, defined as a response being placed within 200 pixels of the non-target location. There was group effect on misbinding errors (*F*(2,51)=25.62, *P*<0.001), the result of PTA+ patients making significantly more misbinding errors than PTA-(*t*(51) =-5.60, *P*<0.001) and controls (*t*(51)=6.65, *P*<0.001; Figure 2D).

We calculated the entropy ratio of the distribution of responses across the x and y axes in the transformed space (Figure 2C). This allowed us to quantify the influence of non-target item on misbinding directly. There was a group effect on the entropy ratio (Figure 2E; *F*(2,51)=9.33, *P*<0.001). PTA+ patients had a significantly higher entropy ratio than PTA-(*t*(51)=-2.26, *P*=0.042) and controls (*t*(51)=4.31, *P*<0.001). There was a group effect on the entropy ratio across a wide range of bin widths (Supplementary Fig.2).

### Impaired binding ability is transient and specific to a period of PTA

Eighteen patients returned for follow-up (average 182 days post-baseline, Supplementary Table 1). Cognitive function generally improved at follow-up (Supplementary Fig.3; Supplementary Table 3), and memory binding in PTA+ patients normalised (Figure 2G). There was a group by visit interaction (*F(1,15)*=20.99, *P*<0.001). This was driven by a reduction in misbinding errors in PTA+ patients between visits but no longitudinal change in PTA-patients (Figure 2H; PTA+ v1 > PTA+ v2 (*t*(7)=4.60, *P*=0.0025); PTA – v1 > PTA-v2 (t(8)=-0.180, *P*=0.862)). There were no significant longitudinal changes in entropy ratio, but PTA+ patients were no longer abnormal compared to PTA-patients (Figure 2H; *t*(8.76)=-0.25, *P*=0.807).

### Patients in post-traumatic amnesia show increased low frequency power

Resting EEG showed changes in power across a range of frequency bands in patients with PTA (delta: *F*(2,35)=5.97, *P*=0.006; alpha: *F*(2,35)=9.05, *P*<0.001; gamma: *F*(2,35)=5.08, *P*=0.012; Figure 3A). In the delta band, these effects were driven by PTA+ patients exhibiting increased power compared to controls (PTA+ > CON: *t*(35)=3.43, *P*=0.005) and a trend towards an increase compared to PTA- (PTA+ > PTA-: *t*(35)=-2.16, *P*=0.057). In the alpha band, PTA+ showed reduced power compared to controls (PTA+ < CON: *t*(35)=-4.25, *P*<0.001) and a trend towards a decrease compared to PTA- (PTA+ < PTA-: *t*(35= 2.09, *P*=0.065). PTA+ patients also showed reductions in beta power (PTA+ < CON: *t*(35)=-3.01, *P*=0.014; PTA- < CON: *t*(35)=-2.66, *P*=0.018). In the gamma band, the PTA- group showed reduced power compared to controls (PTA- < CON: *t*(35)=-2.97, *P*=0.016), with no differences in the PTA+ group.

**Figure 3.**
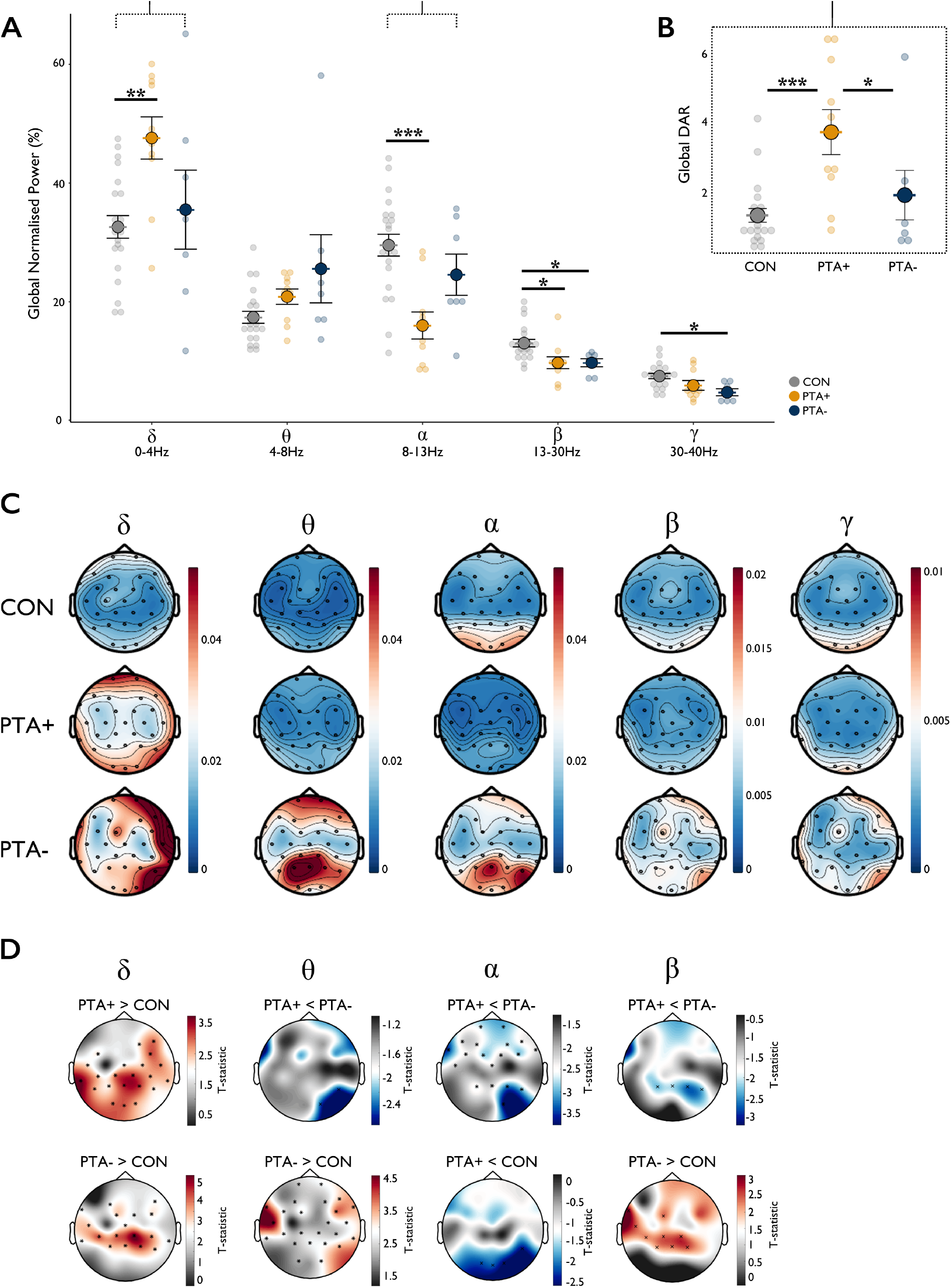
Global normalised power in healthy controls, PTA+ and PTA- TBI patients at baseline. **(A)** Mean global normalised power for all groups across all frequency bands. **(B)** Global delta to alpha ratio across all groups. **(C)** Topoplots depicting mean distribution of power for each group in each frequency band. Blue denotes low power, red denotes higher power. **(D)** Cluster based statistical comparisons of power in (left to right) delta, theta, alpha and beta frequency bands between PTA+, PTA- and healthy controls.

Visual inspection of topoplots (Figure 3C) indicated abnormal patterns of power across multiple frequency bands in both TBI groups. In the delta band, direct comparison between patient groups and controls showed increased power for both PTA+ (*P*=0.002) and PTA-patients in frontal, parietal, and temporal channels, with PTA+ showing a more widespread cluster extending into occipital channels. Conversely, for alpha, PTA+ showed reduced occipital and right parietal power compared to controls, and widespread reductions compared to PTA- in frontal, temporal, and parietal channels. In the theta band, PTA- showed increased power compared to controls in temporal and parietal channels and increased power compared to PTA+ in a right parietal-occipital cluster of channels. Changes in the beta band were of a similar pattern to theta. The PTA- group showed increased power in temporal and parietal channels compared to controls (*P*=0.023) and increased parietal power compared to PTA+ (*P*=0.004). There were no clusters found between any of the groups in the gamma frequency band.

EEG abnormalities in the PTA+ group were transient, and power had normalised at follow-up (Figure 4A). There was a significant effect of visit in delta and alpha (delta: *F*(1,8) =13.65, *P*=0.006; alpha: *F*(1,8)=8.37, *P*=0.020). This was the result of increases in alpha and decreases in delta in the PTA+ group between baseline and follow-up (alpha: PTA+v1 < PTA+v2 (*t*(4)=-4.0726, *P*=0.030); delta: PTA+v1 > PTA+v2 (*t*(4)=4.1234, *P*=0.029)). There was also a significant group by time interaction in beta and gamma but no longitudinal effects were observed in theta (beta: *F*(1,8)=13.23, *P*=0.007; gamma: *F*(1,8)=6.30, *P*=0.036). Figure 4C depicts the spatial distribution in channel space of the change across time (follow-up minus baseline) for each frequency band for PTA+ and PTA-patients.

**Figure 4.**
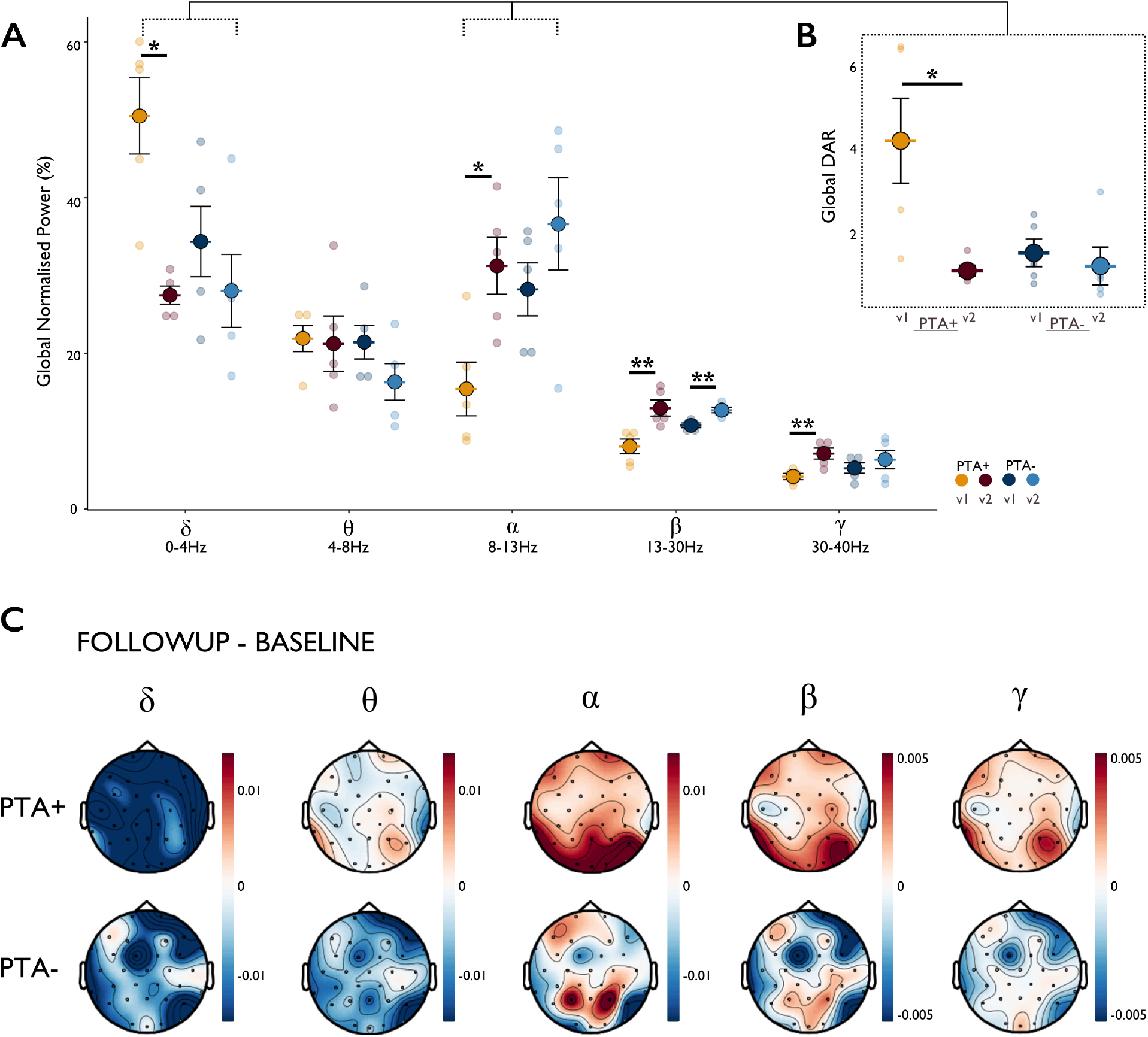
Global normalised power in PTA+ patients and PTA- TBI patients at follow-up. **(A)** Mean global normalised power for all patients with baseline and follow-up across all frequency bands. **(B)** Global delta to alpha ratio. **(C)** Topoplots showing change between baseline and follow-up (follow-up minus baseline) in PTA+ and PTA- TBI patients in power across all frequency bands. Blue denotes absolute reduction in power at follow-up from baseline, white denotes no change, red denotes absolute increase in power at follow-up from baseline.

EEG differences between PTA+ and PTA- were particularly marked in delta and alpha bands, so the delta/alpha ratio (DAR) was calculated. DAR has been used as a sensitive marker of cerebral dysfunction.^32–34^ Group differences in global DAR were present (Figure 3B; *F*(2,35)=9.12, *P*<0.001), the result of significantly higher in PTA+ compared to both PTA- (*t*(35)=-2.51, *P*=0.025) and controls (*t*(35)=4.26, *P*<0.001). Abnormalities in DAR normalised at follow-up, as demonstrated by a significant group by time interaction (*F*(1,8)=5.48, *P*=0.047), the result of a reduction in DAR in PTA+ with no change in PTA- group (PTA+v1 > PTA+v2: *t*(4)=3.008, *P*=0.040; Figure 4B).

### Individual case studies

To better describe the transient binding impairment and how this might relate to the transient shift towards slow wave power, we considered these changes at the single patient level. Figure 5 illustrates four individual case studies to highlight that the EEG changes reported here are more sensitive to abnormalities occurring during a period of PTA than conventional routine clinical imaging. Case studies one to three show TBI patients during a period of PTA at baseline and at follow-up once they were no longer in PTA. Case study four shows a TBI patient who was not in PTA.

**Figure 5.**
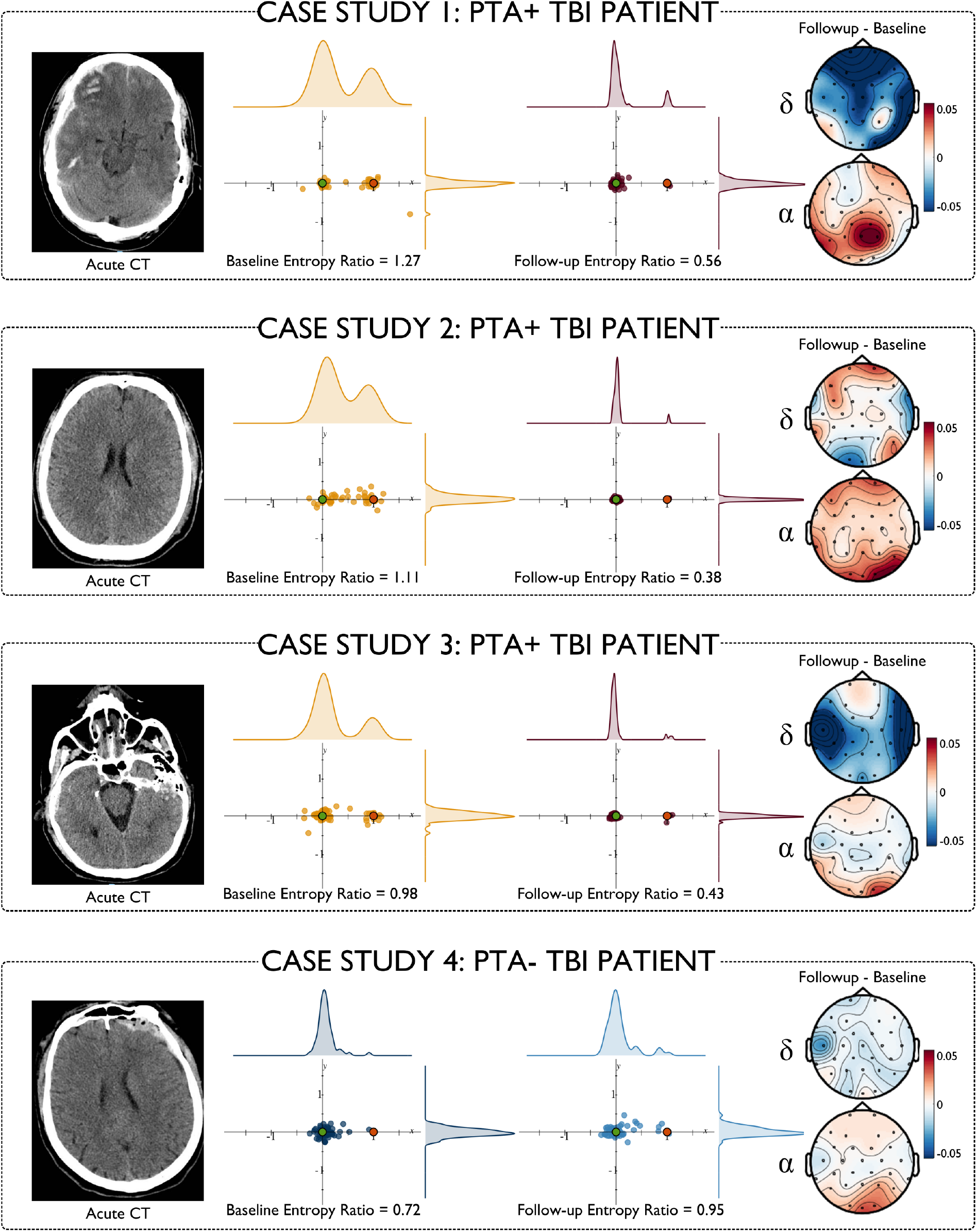
Individual case studies. CT head scans at admission are shown on the far left of each case study panel. The distribution of responses in correctly identified trials in the precision spatial working memory task are shown for baseline and follow-up in the middle panels. Topoplots (right) depict changes in delta (top) and alpha (bottom) between baseline and follow-up.

Case study one is a male in his 40s with a moderate-severe TBI acquired through a fall from standing. Clinical imaging reported presence of subdural haemorrhage (SDH), subarachnoid haemorrhage (SAH), bi-frontal contusions and midline shift. On the day of assessment (day eight post-injury, WPTAS score 8) he was clinically deemed to be in PTA and had a total PTA duration of 12 days. At baseline he showed a bias towards the non-target on the precision working memory task with an entropy ratio of 1.27. He showed a dramatic improvement at follow-up to an entropy ratio of 0.56. At baseline he had a global DAR of 6.46 which reduced to 0.99 at follow-up.

Case study two is male in his 20s with a moderate-severe TBI acquired through a cycling collision. There was no intracranial haemorrhage or space-occupying lesions on the initial clinical imaging. Further imaging with MRI revealed evidence of diffuse axonal injury (DAI). He was assessed on day 24 post-injury, scoring 7 on the WPTAS and thus still in a period of PTA. He had a total PTA duration of 38 days. Working memory binding performance at baseline was poor with an entropy ratio of 1.11 which reduced to 0.38 at follow-up. At baseline his global DAR was 6.52 and reduced to 0.82 at follow-up.

Case study three, a male in his 60s, acquired a moderate-severe TBI acquired while cycling. He had a total PTA duration of 28 days. Clinical imaging showed a left SDH, left temporal contusions, (probable) right extradural haemorrhage (EDH), skull base fractures and pneumocephalus. On assessment he was clinically deemed to be in PTA (WPTAS score 8; day 5 post-injury). At baseline he had an entropy ratio of 0.98 which improved to 0.43 at follow-up. His global DAR was 4.21 at baseline and reduced to 0.75 at follow-up.

Case study four is a male in his 30s with a moderate-severe TBI acquired through a road accident as a pedestrian. Clinical imaging reported left-sided extra-axial haematoma with associated comminuted fracture involving the left frontal bone. Evidence of DAI was present on MRI. He had a PTA duration of 0 days and was thus not in PTA at the time of assessment (WPTAS score 12; day 5 post-injury). At baseline he did not demonstrate any binding deficit, with an entropy ratio of 0.72 which increased slightly at follow-up to 0.95. At baseline he had a global DAR of 2.04 which decreased to 0.48 at follow-up.

### Power abnormalities in acute TBI are associated with working memory performance

To quantify the relationship between EEG measures and working memory performance following TBI, we grouped PTA+ and PTA- patients to examine the relationship between a shift towards low frequency power and cognitive performance (Figure 6). Global delta power was associated with working memory performance in TBI patients but not controls (Figure 6A; Misbinding errors (TBI: *R*=0.52, *P*=0.038; Controls: *R*=-0.07, *P*=0.780); Paired associates learning (TBI: *R*=-0.56, *P*=0.018; Controls: *R*=0.24, *P*=0.38)). Global alpha power was positively associated with performance on the paired associates learning task in TBI patients but showed a negative relationship to performance in controls (TBI: *R*=0.74, *P*<0.001; Controls: *R*=-0.58, *P*=0.025). Across TBI patients, global DAR correlated with the entropy ratio of response distribution for the precision working memory task (Figure 6B; TBI: *R*=0.51, *P*=0.04; controls: *R*=-0.29, *P*=0.22); Misbinding errors (Figure 6C; TBI: *R*=0.34, *P*=0.02; controls: *R*=-0.23, *P*=0.33); Paired Associates Learning (Figure 6D; TBI: *R*=-0.6, *P*=0.01; controls: *R*=0.44, *P*=0.10). The total duration of PTA, a proxy of injury severity, was not associated with power in either delta or alpha frequency bands nor in the DAR.

**Figure 6.**
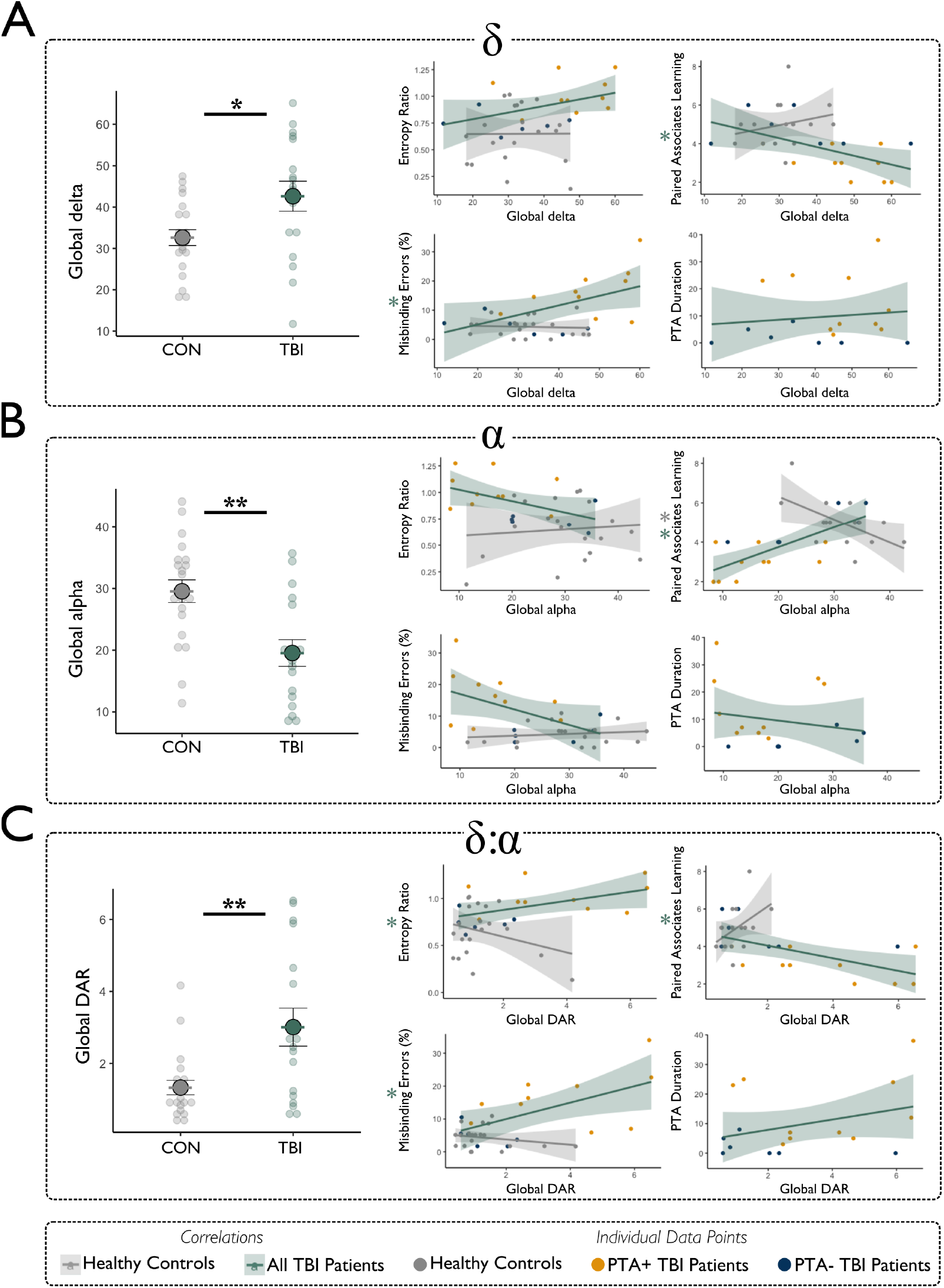
Relationship between global normalised power and cognitive impairment. **(A)** Global delta power was increased in the TBI group compared to healthy controls and correlated with proportion of misbinding errors and performance on the paired associates learning task. **(B)** Global alpha was significantly decreased in TBI patients compared to healthy controls, and significantly correlated with the performance on the paired associates learning task. **(C)** Global DAR was significantly increased in TBI patients compared to healthy controls and correlated with the entropy ratio, proportion of misbinding errors and performance on the paired associates learning task.

### TBI patients demonstrate theta hyperconnectivity and alpha hypoconnectivity

Next, we assessed connectivity within the delta, theta, and alpha bands. Phase synchronisation was quantified using the dwPLI. Whole brain connectivity matrices were constructed on a channel-wise basis for controls and patients in each band separately (Figure 7). Network based statistics revealed that patients show theta hyperconnectivity compared to controls across one robust network consisting of 19 edges and 15 nodes (*P*=0.005; Figure 7B). In the alpha band there was a single robust network of hypoconnectivity in TBI patients compared to controls consisting of 6 edges and 6 nodes (*P*=0.020; Figure 7C). There was a relationship between the mean dwPLI across the theta network and the total duration of PTA (*R*=0.57, *P*=0.016). The mean dwPLI in the alpha network did not show a relationship with PTA duration (*R*=-0.08, *P*=0.760). At follow-up, whole-brain connectivity in the delta and theta bands decreased and increased in the alpha band (Supplementary Fig.4).

**Figure 7.**
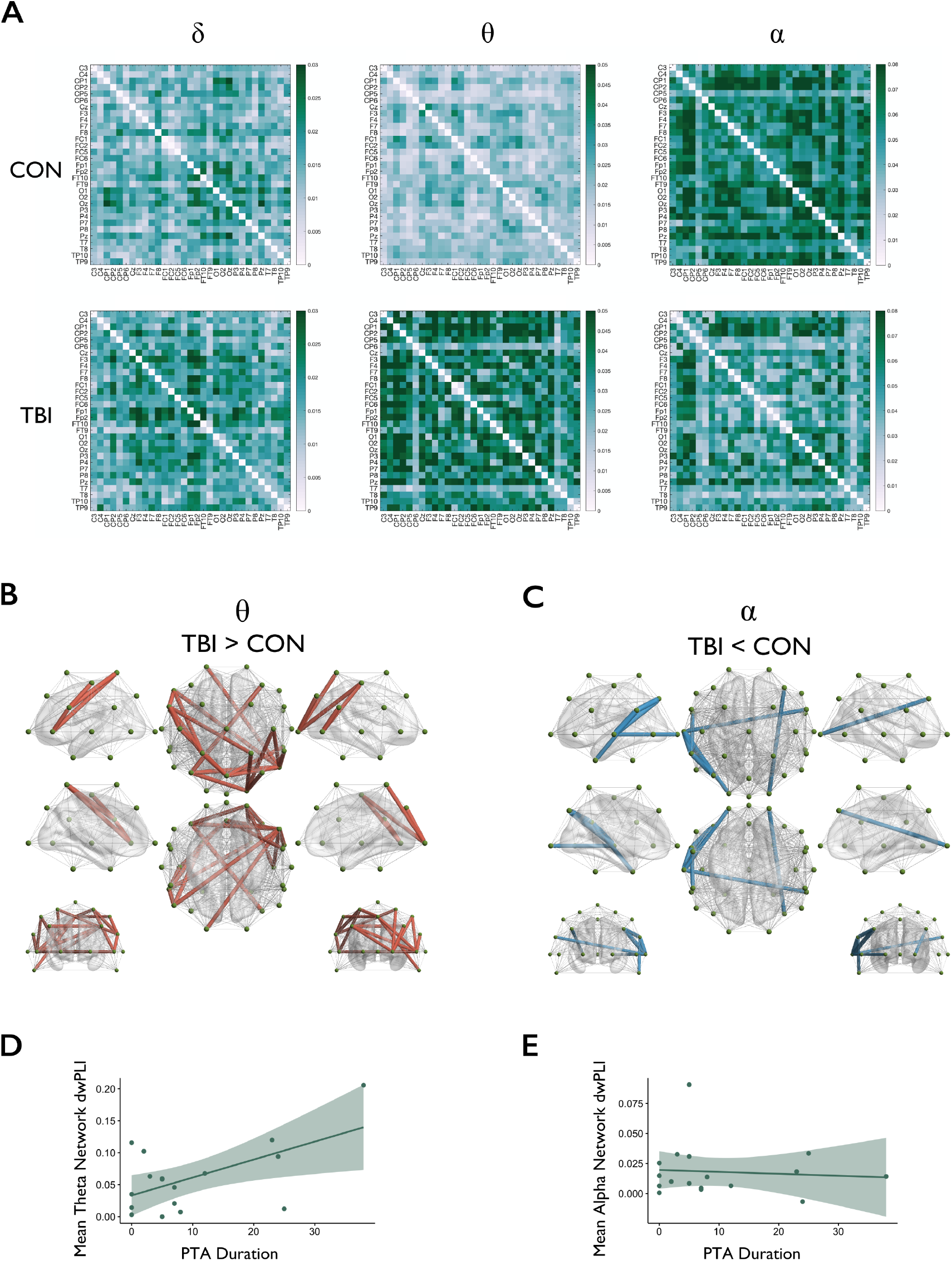
Phase synchronisation. **(A)** Whole brain connectivity matrices in delta, theta and alpha frequency bands for healthy controls (top row) and all TBI patients (bottom row). **(B)** Network based statistics revealed one robust network of hyperconnectivity in TBI patients compared to controls in the theta band and **(C)** one robust network of hypoconnectivity in the alpha band. **(D)** Mean dwPLI in the resulting theta network was positively associated with total PTA duration **(E)** Mean dwpLI in the resulting alpha network showed no relationship to total PTA duration.

### Theta-gamma cross-frequency coupling is not altered in acute TBI

Phase-amplitude coupling, specifically frontal theta phase to parietal and temporal gamma amplitude, was quantified using the MI. Contrary to our hypothesis, patients did not show increased phase-amplitude coupling for either frontal to parietal (t(35.97)=-1.413, *P*=0.1662) or frontal-temporal (*t*(34.42)=-0.634, *P*=0.5303) modulation. There were no significant longitudinal changes in patients returning for follow-up in the mean modulation index for either frontal-parietal (*t*(9)=1.567, *P*=0.1516) or frontal-temporal (*t*(9)=0.276, *P*=0.7891) channels (Supplementary Fig.5).

## Discussion

Post-traumatic amnesia (PTA) is a common consequence of TBI characterised by a profound but transient cognitive disturbance. Here, we show that PTA is associated with a marked impairment in the binding of information in working memory that resolves when patients emerge from PTA. This behavioural abnormality is associated with a shift to low-frequency oscillations on EEG, quantified by an increase in delta-alpha ratio of EEG power (DAR), which is specifically seen in PTA and not TBI patients without PTA. Increased DAR correlated with memory binding failures, potentially indicating a causal role in the disruption of working memory function. Connectivity is also non-specifically increased in the theta band and decreased in the alpha band following TBI. The results suggest that abnormal low-frequency oscillations, which are seen in many disease states, may disrupt the precise synchronisation of neural oscillations that are necessary for working memory function.

We used a precision spatial working memory task to assess working memory function. This has several important advantages. Most importantly, distinct types of errors can be accurately assessed. Subjects were required to accurately encode both the identity of an object and its spatial location in working memory. Separate object identification and location phases probed identity and spatial information. Importantly, subjects in PTA had an accurate memory of object identity and spatial location on the majority of trials. However, when objects were successfully identified, there were high levels of misbinding i.e., fractals on the screen were moved to the incorrect spatial location. This error was not random. Rather, patients in PTA systematically chose the location of the non-target fractal indicating that both spatial information and object identity had been encoded, but that the binding of this information was impaired.

Misbinding within working memory has generally been assessed using simple measures of the distance between a response and the target or non-target location. This approach relies on an arbitrarily defined threshold to decide whether a response is within a specific location and does not fully describe the distribution of responses across trials. This approach limited our ability to assess the relationship between EEG abnormalities and working memory function, so we developed a novel approach to quantifying the distribution of responses across trials. An entropy ratio was calculated that quantified the relationship between the distribution of responses across the x and y axes in a normalised space, where the target and non-target are transformed to be in a consistent location relative to one another (Figure. 1). Entropy in this context quantifies the probability that a response will occur at a particular location along either the x or y axis that defines the 2D response space. Low entropy indicates a high probability for the clustering of responses around a particular spatial location e.g., around the target location for correct responses. Increasing entropy indicates a greater probability that responses have occurred at other locations. Calculating separate entropies for x and y axes allows the influence of the distractor location to be separated from the presence of random responses because misbinding errors will increase entropy only in the x axis, whereas random responses would increase entropy in both dimensions.

Our entropy results clearly show that the spatial errors in PTA are non-random. The errors cluster around the location of the distractor, resulting in an increased entropy ratio (x/y). This shows that subjects are engaged in the task, encode an object’s identity and location but show impairments of working memory binding. This approach is informative when applied to individual cases, as we have shown. It is informative to display responses in a transformed space as this visualisation can be used to illustrate the presence of misbinding errors and the absence of random responses. Entropy ratios are increased for patients with PTA and normalise in the same patients after their emergence from PTA. Hence, we show that a precision working memory task of this type could be used to quantify the presence of PTA and the misbinding measures are likely to be a sensitive measure of how a PTA state changes over time. A similar approach has recently been released as a mixture modelling toolbox designed for distinguishing the sources of spatial memory error.^71^ This approach estimates the proportion of non-target, target, and guess responses by fitting probability density functions to the data in a 2D space. Like our approach, this allows for a threshold-free analysis of spatial data but calculates different summary statistics. The entropy ratio we present provides a complementary way to describe the distractor influence and random error with a single continuous measure.

Resting state EEG measures were then used to identify electrophysiological abnormalities in acute TBI patients and controls. In TBI, a shift towards slower wave power has previously been associated with long-term neuropsychological outcome, functional outcome and clinical symptoms of TBI.^30,72^ EEG slowing is often viewed as a nonspecific sign of cerebral dysfunction. We show that patients in post-traumatic amnesia show decreased alpha power, increased delta power and a significantly higher DAR than TBI patients without PTA and controls. This abnormality correlated with the misbinding of information in working memory and normalised at follow-up. This suggests that increased DAR may be a sensitive electrophysiological marker of PTA that is closely related to the key cognitive impairments seen in this state.

Low frequency oscillations are observed in a range disease states.^27–29^ For example, patients with Alzheimer’s Disease (AD) often show a shift towards increased low frequency oscillations characterised by delta synchronization and alpha de-synchronisation.^8,11,73^ Early working memory abnormalities are seen in AD and increased delta power is associated with working memory impairments.^74^ In addition, increased low frequency oscillations are also associated reduced levels of awareness.^27^ This can be studied in animal models and we have previously used voltage imaging in rodents to show that anaesthesia is associated with low frequency hypersynchrony that is associated with reduced communication between cortical regions. ^75,76^ Hence, increased low-frequency oscillations are common to states of disordered consciousness, dementia and PTA and may provide a common electrophysiological mechanism for the disruption of the cortical dynamics that are necessary for the disruption of higher cognitive functions including working memory and awareness. Patients in PTA provide a unique way to study transient changes in relationship between electrophysiological abnormalities and working memory binding. Here, we show that DAR normalizes in conjunction with the emergence from PTA and the normalisation of working memory binding impairments. Taken together, our results suggest that the presence of increased low-frequency oscillations may disrupt electrophysiological interactions necessary for successful integration of object and location memory, and this mechanism may be relevant across a range of disease states.

To investigate how low frequency oscillations might disrupt working memory, we explored the effect of TBI on cortical connectivity. Co-ordinated activity across the brain is important for cortical communication and we calculated the phase lag index (dwPLI), which quantifies correlation between different EEG channels. Acute TBI patients showed theta hyperconnectivity and alpha hypoconnectivity using this measure, but these changes were not specific to PTA. In addition, although theta connectivity between frontal and temporal-parietal regions has been shown to increase with working memory load and executive control,^16^ it did not correlate with working memory performance following TBI. However, theta connectivity increased with PTA duration, suggesting it might relate to injury severity^77^. In addition, we explored phase-amplitude coupling in the theta/gamma bands, as this has been implicated in working memory binding.^25,26^ This was not abnormal following TBI and did not relate to working memory performance. One explanation for this null result is that changes in phase-amplitude coupling might only be seen during working memory performance. As we only investigated resting state EEG data, we cannot explore the relationship between neural oscillatory dynamics and distinct working memory stages. Future work could usefully explore the relationship between EEG abnormalities and working memory impairment during PTA event related analyses of task performance.

As expected, our patients were heterogenous in terms of TBI pathophysiology. The pattern of TBI seen on neuroimaging varied but did not explain the presence or absence of PTA. Nevertheless, future work in a larger sample might investigate the relationship between distinct types of brain injury and the EEG/behavioural abnormalities we have observed. In addition, there was also a trend towards the PTA group being slightly older than controls. This is unlikely to explain our results. During healthy ageing there is a linear decrease of slow frequency resting-state activity.^78^ This would suggest that if age alone were influencing the results, then the PTA+ group would be expected to show lower delta power than controls. In fact, the opposite was the case, and PTA+ showed significantly greater delta power than controls. The trend towards a difference in age between the groups should therefore not alter the interpretation of a significant shift towards lower frequency oscillations during PTA.

In summary, acute TBI patients in a period of PTA show a marked impairment of working memory binding. We quantified this using a novel entropy ratio measure and showed how this relates to electrophysiological changes on EEG. Our results demonstrate a clear relationship between a shift to pathological oscillatory slowing and transient working memory impairment in PTA, which is informative in understanding the profound but transient effects of TBI on higher cognitive functions.

## Data Availability

Data are available upon reasonable request to the authors.

## Abbreviations

CON: healthy control
DAI: diffuse axonal injury
DAR: delta to alpha ratio
DR: delayed recall
dwPLI: debiased weighted phase lag index
FDR: false-discovery rate
FFT: fast fourier transform
GCS: Glasgow coma scale
ICA: independent component analysis
IR: immediate recall
LOC: loss of consciousness
MARA: multiple artifact rejection algorithm
MI: modulation index
MTW: major trauma ward
NBS: network based statistic
PAC: phase amplitude coupling
PAL: paired associates learning
PTA: post-traumatic amnesia
SAH: subarachnoid haemorrhage
SD: standard deviation
SDH: subdural haemorrhage
SEM: standard error of the mean
TBI: traumatic brain injury
WPTAS: Westmead post-traumatic amnesia scale

## Acknowledgements

The authors would like to thank all the participants who took part in this study. The authors also thank the staff at the Major Trauma Ward and the Neurosurgery, Emergency and Trauma Research Team, St Mary’s Hospital, Imperial NHS Healthcare Trust, London, UK for their assistance in patient screening.

## Funding

The work was supported by an Academy of Medical Sciences Starter Grant for Clinical Lecturers (awarded to NG) and by an equipment grant from the Department of Medicine, Imperial College London. E-JM is supported by the UK Dementia Research Institute Care Research and Technology Centre. GS is supported by NIHR. DJS is supported by the UK Dementia Research Institute Care Research and Technology Centre, an NIHR Professorship (NIHR-RP-011-048), the NIHR Clinical Research Facility and Biomedical Research Centre at Imperial College Healthcare NHS Trust & The Royal British Legion Centre for Blast Injury Studies.

## Competing interests

The authors report no competing interests.

## Notes

### Competing Interest Statement

The authors have declared no competing interest.

### Author Declarations

The study was approved by the West London Research Ethics Committee (09/HO707/82).

