## Supplementary for "Pathological slow-wave activity and impaired working memory binding in post-traumatic amnesia"

### Supplementary Materials

**Supplementary Figure 1. Neuropsychological performance at baseline**

**Supplementary Figure 2. Entropy ratio at different bin widths**

**Supplementary Figure 3. Neuropsychological performance at follow-up**

**Supplementary Figure 4. Phase synchronisation at follow-up**

**Supplementary Figure 5. Phase-amplitude coupling at baseline and follow-up**

**Supplementary Table 1. Detailed clinical characteristics**

**Supplementary Table 2. Neuropsychology at baseline**

**Supplementary Table 3. Neuropsychology at follow-up**

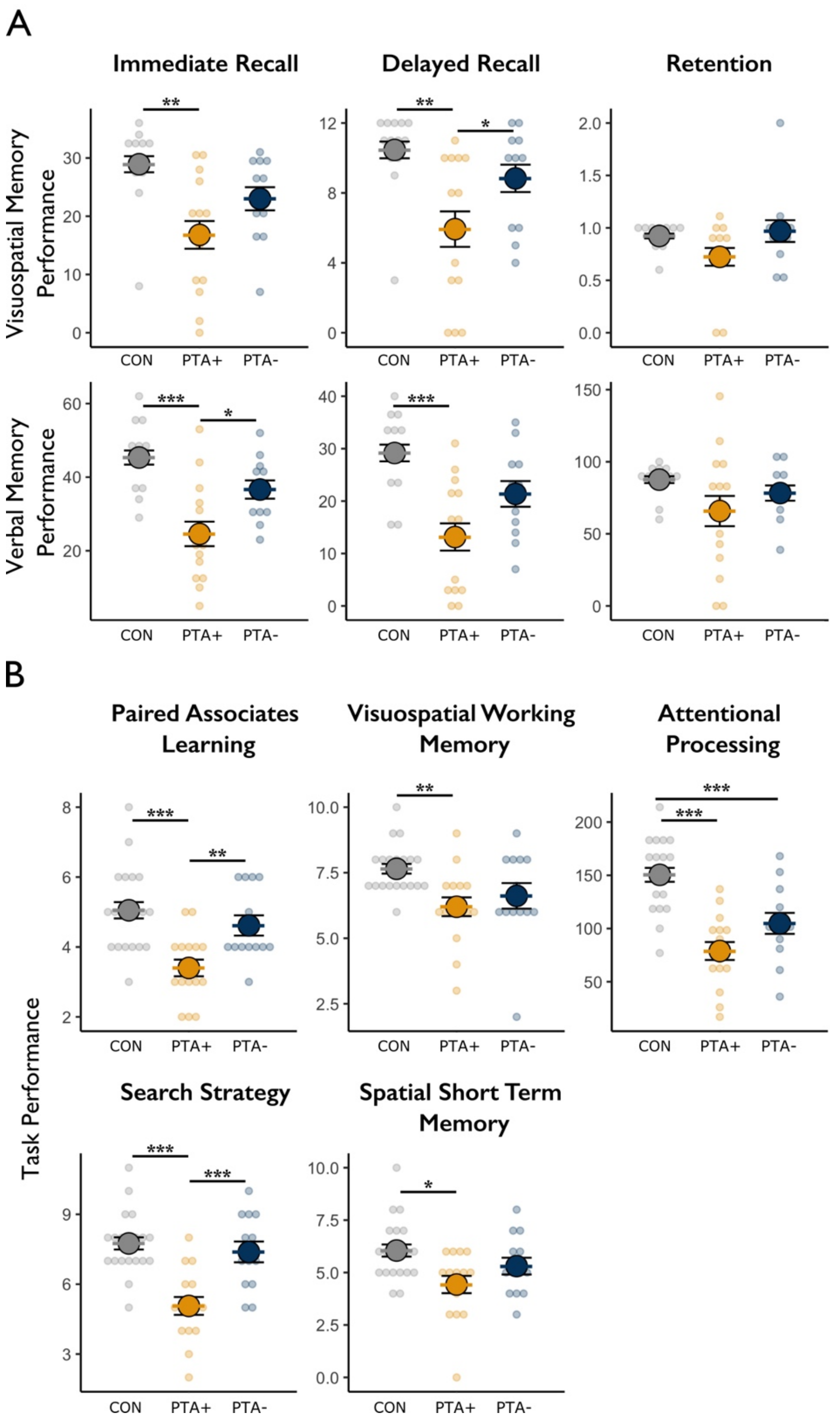


**Supplementary Figure 1. Neuropsychological performance of PTA+ and PTA- TBI patients and healthy controls at baseline.** (A) Visuospatial memory performance as measured using the BVMT (top row) and verbal memory performance as measured using the Logical Memory test (bottom row) for immediate recall, delayed recall, and retention. (B) Performance on the computerised battery of tests on the paired associates learning, visuospatial working memory, attentional processing, search strategy and spatial short term memory tasks.

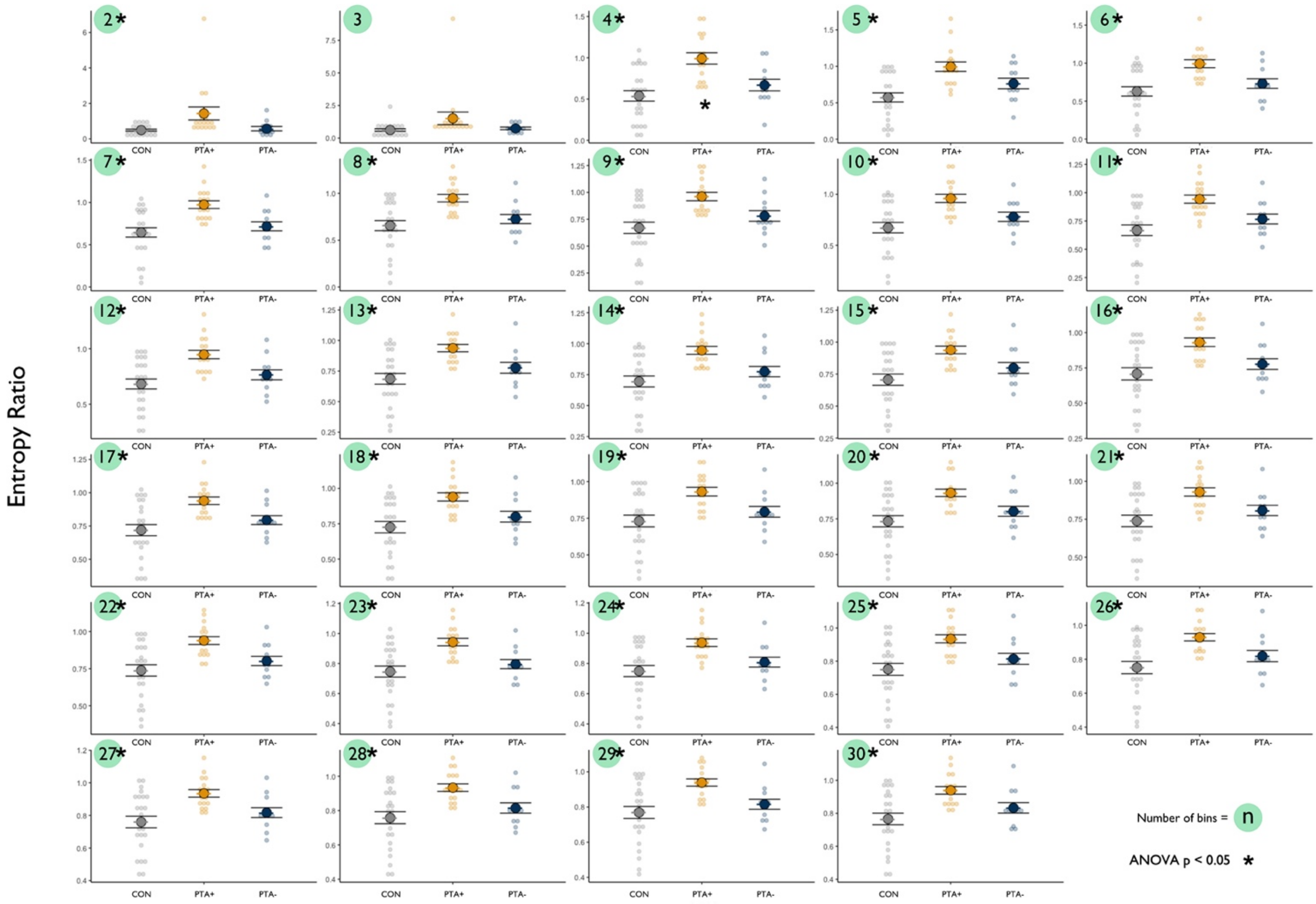


**Supplementary Figure 2. Entropy ratio at different bin widths.** Entropy ratio for healthy controls, PTA+ and PTA- TBI patients calculated using n=2 to n=30 bins. There was a robust effect of group when using a different number of bins to calculate the entropy ratio with the exception of n=3 bins.

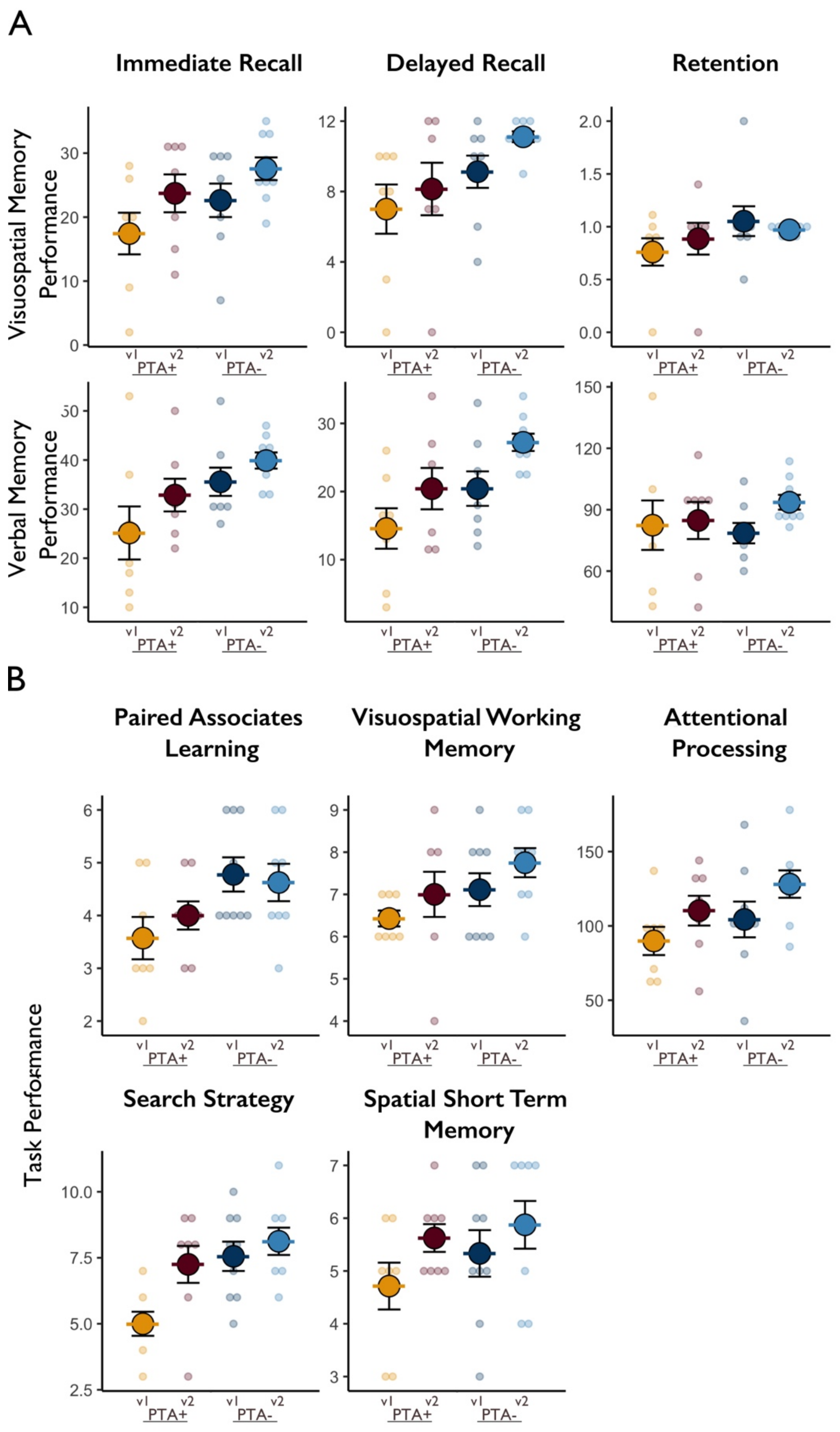


**Supplementary Figure 3. Neuropsychological performance of PTA+ and PTA- TBI patients who returned for follow-up at visit 1 and visit 2. (A)** Visuospatial memory performance as measured using the BVMT (top row) and verbal memory performance as measured using the Logical Memory test (bottom row) for immediate recall, delayed recall, and retention. **(B)** Performance on the computerised battery of tests on the paired associates learning, visuospatial working memory, attentional processing, search strategy and spatial short term memory tasks.

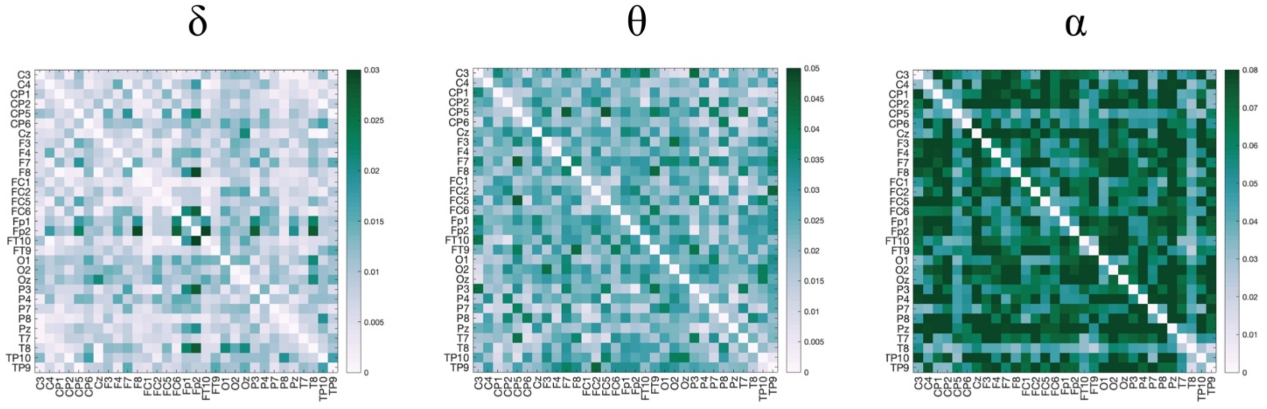


**Supplementary Figure 4. Phase synchronisation at follow-up** in the delta, theta and alpha frequency bands (from left to right).

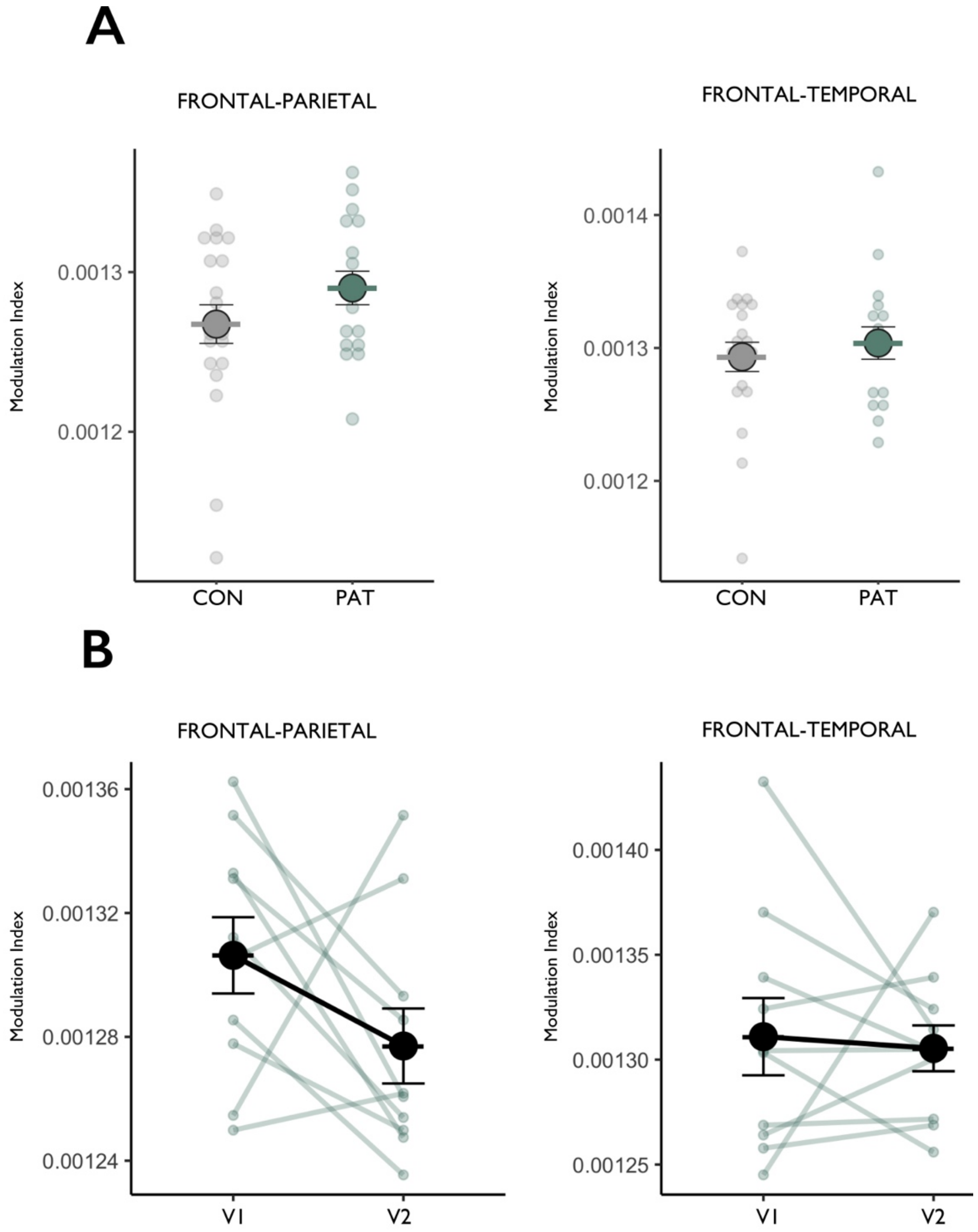


**Supplementary Figure 5. Frontal theta phase to parietal and temporal gamma amplitude coupling. (A)** Average modulation index between frontal and parietal channels (left) and frontal and temporal channels (right) in patients and controls at baseline. **(B)** Longitudinal changes in phase-amplitude coupling between frontal-parietal (left) and frontal-temporal (right) channels in patients at follow-up. Error bars represent standard error of the mean.

**Supplementary Table 1 Detailed clinical characteristics**

| PTA Status | WM task | EEG | V2 | Age Range | Sex | Time since injury (days) | Cause of Injury | PTA Duration (days) | Lowest GCS | Length of Hospital Admission (days) | Clinical Imaging Report | Medications |
|---|---|---|---|---|---|---|---|---|---|---|---|---|
| PTA- | Yes | No | Yes | 40-44 | M | 11 | Assault | 10 | 13 | 11 | SAH; anterior temporal lobe contusion; parietal and occipital skull fracture; thrombosed venous sinus | Ondanestron 4mg; Lactulose 14ml; Levetiracetam 1g; Paracetamol 1g; Senna 15mg; Morphine 10mg |
| PTA+ | Yes | No | No | >20 | F | 7 | Sports injury: Football | 2 | 3 | 8 | Negative | NA |
| PTA+ | Yes | No | Yes | 45-49 | M | 6 | RTA: Cyclist | 2 | 15 | 6 | SAH | NA |
| PTA+ | Yes | No | Yes | 20-24 | M | 21 | Fall from height | 22 | 8 | 21 | SDH; SAH; ME; MS; | NA |
| PTA+ | Yes | No | Yes | 35-39 | M | 6 | RTA: Ped vs Car | 23 | 3 | 23 | SDH; EAH | NA |
| PTA- | Yes | No | Yes | 30-34 | M | 3 | Assault | 0 | 14 | 5 | SAH | Ciprofloxacin; Dihydrocodeine; Morphine; Ondanestron; Senna |
| PTA- | Yes | No | No | 20-24 | M | 26 | Assault | 28 | 3 | 26 | EDH; ME; MS; SDH; SAH | Paracetamol 1g; Tinzaparin; Lansoprazole 30mg; Ibuprofen 400mg; |
| PTA- | Yes | No | No | 60-64 | M | 2 | Fall from standing | 2 | 14 | 2 | SDH; SAH | Ondanestron 4mg; Levetiracetam 1g; Dihydrocodeine 30mg; Paracetamol 1g; |
| PTA- | Yes | No | Yes | 40-44 | M | 2 | Fall from height | 0 | 8 | 2 | SAH | Dihydrocodeine 30mg; Paracetamol 1g |
| PTA+ | Yes | No | No | 50-54 | M | 5 | RTA: Cyclist | 6 | 14 | 6 | SAH | Senna, Dihydrocodeine, Macrogal, morphine, levetiracetam, ondanestron |
| PTA+ | Yes | No | No | 45-49 | M | 32 | Assault | 30 | 10 | 30 | SDH; SAH | Haloperidol 1mg; Lansoprazole 30mg; Lorazepam 0.5mg; Morphine 2.5mg; Paracetamol 1g; Zopiclone 7.5mg |
| PTA- | Yes | No | Yes | 60-64 | F | 3 | Fall from height | 0 | 8 | 2 | Small temporal contusion + SAH; SDH (bifrontal) | Citalopram |
| PTA+ | Yes | No | No | 40-44 | M | 16 | RTA: Motorcyclist | 20 | 3 | 30 | SAH; DAI | NA |
| PTA- | Yes | Yes | Yes | 30-34 | M | 5 | RTA: Ped vs Car | 0 | 14 | 21 | DAI | NK |
| PTA+ | Yes | Yes | No | 40-44 | M | 1 | Fall from height | 5 | 3 | 5 | SAH; inferior frontal intraparenchymal haemorrhage | Chlordiazepoxide (10mg); Polyethylene glycol with electrolytes |
| PTA+ | Yes | Yes | No | 60-64 | F | 3 | RTA: Ped vs Bus | 5 | 15 | 5 | SAH; SDH; occipital skull fracture; left frontal contusion | Morphine sulphate 20mg; ondansetron 4mh; dihydrocodeine 60mg; paracetamol 1g; phenytoin 300mg; chloraphenamine 4mg; lactulose 15ml; senna 15mg |
| PTA+ | Yes | Yes | No | 55-59 | M | 16 | Assault | 23 | 6 | 23 | SAH | Haliperidol 2mg; Oxycodon 5mg; Docusate 200mg; Senna 10mg; Paracetamol 1g; Salbutamol 2.5mg; Chlorphenamine 4mg; Lansoprazole 30mg; Nicotine 1 patch; Nystatin 100000 units; |
| PTA+ | Yes | Yes | Yes | 45-49 | M | 8 | Fall from standing | 12 | 14 | 12 | SDH; SAH; bifrontal contusions; midline shift | Morphine sulphate 20mg; cyclizine 50mg; dihydrocodine 60mg; paracetamol 1g; carbimazole 5mg; ondansetron 8mg; metformin 500mg; lactulose 10ml; levetiracetam 500mg; |

| | | | | | | | | | | | | |
|---|---|---|---|---|---|---|---|---|---|---|---|---|
| PTA+ | Yes | Yes | Yes | 25-29 | M | 20 | RTA: Ped vs Car | 25 | 3 | 30 | SDH | Dihydrocodeine 30mg; ondansetron 4mg; haloperidol 3mg; docusate 200mg; paracetamol 1g; flucloxacillin 1g; chlordiazepozide 20mg; senna 7.5mg; Enoxaparin 40mg |
| PTA- | No | Yes | No | 40-44 | F | 3 | RTA: Cyclist | 0 | 14 | 6 | SAH; parenchymal contusions | NK |
| PTA+ | Yes | Yes | Yes | 25-29 | M | 24 | RTA: Cyclist | 38 | 7 | 37 | DAI | Docusate 200mg; Senna 10ml; Enoxaparin 40mg; Flucloxacillin 1g; Haloperidol 2.5mg; Paracetamol 1mg; Ciprofloxacin 400mg; Levetiracetam 1000mg |
| PTA+ | Yes | Yes | No | >20 | M | 7 | Fall from height | 24 | 7 | 9 | Left temporal contusion; pneumocephalus; skull fracture | Ondansetron 4mg; Noradrenaline 50ml; Docusate 200mg; Haloperidol 2.5mg; Paracetamol 1g; Co-amoxiclav 625mg; Levetiracetam 1g; Senna 15mg; Sodium chloride nebulised 5ml; |
| PTA+ | Yes | Yes | Yes | 65-69 | M | 5 | RTA: Cyclist | 7 | 13 | 28 | SDH; left temporal contusions; EDH; skull base fractures; pneumocephalus | Lactulose 10ml; levetiracetam 1g; paracetamol 1g; senna 15mg; cyclizine 50mg; morphine sulphate 2mg; naloxine 100 micrograms; ondansetron 4mg; gabapentin 600mg; beclometasone 1 puff; enoxaparin 40mg; ferrous fumarate 210 mg; |
| PTA+ | Yes | Yes | Yes | 50-54 | M | 2 | Sports injury: Ice skating | 3 | 13 | 3 | SDH; SAH; midline shift; right frontal contusion; right temporal contusion; skull fracture | Cyclizine 50mg; paracetamol 1g; levetiracetam 1g; ondansetron 4mg; dihydrocodeine 16mg |
| PTA+ | Yes | Yes | No | 30-34 | M | 8 | Fall from height | 7 | 13 | 9 | EDH; SDH | NK |
| PTA- | Yes | Yes | No | 70-74 | F | 3 | Fall from standing | 0 | 15 | 3 | SDH | NK |
| PTA- | Yes | Yes | Yes | 45-49 | M | 4 | Fall from standing | 0 | 14 | 4 | Contusions in left frontal lobe and right temporal lobe compatible with contrecoup injury; SAH | Dihydrocodeine 30mg; Ondanestron 4mg; Paracetemol 1g; Senna 15mg; Prochlorperazine 10mg |
| PTA- | Yes | Yes | Yes | 20-24 | M | 18 | Assault | 8 | 8 | 9 | Bilateral extra-axial haemorrhages in temporal lobes, SAH, left temporal bone fracture. | Nil |
| PTA- | Yes | Yes | Yes | 20-24 | M | 19 | RTA: Cyclist | 2 | 14 | 2 | DAI | Dihydrocodeine 30mg |
| PTA- | Yes | Yes | Yes | 25-29 | M | 14 | RTA: Motorcyclist | 5 | 14 | 14 | SDH | Paracetamol 1g |

V2= returned for follow-up; M = male; F = female; RTA = road traffic accident; Ped = pedestrian; SAH = subarachnoid haemorrhage; SDH = subdural haemorrhage; ME = mass effect; MS = midline shift; EDH = extradural haemorrhage; DAI = diffuse axonal injury

**Supplementary Table 2. Neuropsychology at baseline**

| Cognitive Domain | Neuropsychological Test | CON Mean (± SD) | PTA+ Mean (± SD) | PTA- Mean (± SD) | ANOVA Group Effect F | p | Post-hoc group differences (FDR corrected) |
|---|---|---|---|---|---|---|---|
| Verbal Memory | Logical Memory Immediate Recall | 45.33 (9.76) | 24.57 (13.74) | 36.64 (8.94) | 11.20 | 0.0002 | PTA+ < CON (p=0.0001)*** PTA+ < PTA- (p=0.0176)* |
| | Logical Memory Delayed Recall | 29.17 (8.12) | 13.14 (10.69) | 21.36 (8.82) | 9.46 | 0.0005 | PTA+ < CON (p=0.0004)*** |
| | Logical Memory Retention | 87.57 (11.93) | 65.79 (43.34) | 78.27 (19.06) | 1.786 | 0.183 | |
| Visuospatial Memory | BVMT Immediate Recall | 28.92 (7.05) | 16.80 (9.81) | 23.00 (7.16) | 7.572 | 0.002 | PTA+ < CON (p=0.0012)** |
| | BVMT Delayed Recall | 10.46 (2.44) | 5.93 (4.18) | 8.83 (2.82) | 6.66 | 0.003 | PTA+ < CON (p=0.0029)** PTA+ < PTA- (p=0.0454)* |
| | BVMT Retention | 0.92 (0.12) | 0.72 (0.35) | 0.97 (0.37) | 2.40 | 0.1058 | |
| | Visuospatial Working Memory | 7.65 (0.93) | 6.20 (1.47) | 6.62 (1.76) | 5.26 | 0.0089 | PTA+ < CON (p=0.0098)** |
| Associative Working Memory | Paired Associates Learning | 5.05 (1.19) | 3.40 (0.99) | 4.62 (1.04) | 10.09 | 0.0002 | PTA+ < CON (p=0.00018)*** PTA+ < PTA- (p=0.00779)** |
| Spatial Short Term Memory | Spatial span | 6.05 (1.47) | 4.43 (1.70) | 5.31 (1.44) | 4.63 | 0.0150 | PTA+ < CON (p=0.0120)* |
| Search Strategy | Self-ordered search | 7.75 (1.33) | 5.07 (1.58) | 7.38 (1.61) | 15.24 | <0.0001 | PTA+ < CON (p<0.0001)*** PTA+ < PTA- (p=0.00025)*** |
| Attentional Processing | Feature Match | 150.45 (33.41) | 78.87 (33.68) | 104.85 (35.51) | 19.48 | <0.0001 | PTA+ < CON (p<0.0001)*** PTA- < CON (p=0.00082)*** |

**Supplementary Table 3. Neuropsychology at follow-up**

| Cognitive Domain | Neuropsychological Test | PTA+ Mean (± SD) | PTA- Mean (± SD) | Mixed Effects Model: Group F | Group p | Timepoint F | Timepoint p | Group x Timepoint F | Group x Timepoint p |
|---|---|---|---|---|---|---|---|---|---|
| Verbal Memory | Logical Memory Immediate Recall | 32.86 (9.41) | 39.89 (4.94) | 3.65 | 0.0785 | 5.32 | 0.0415* | 0.08 | 0.7786 |
| | Logical Memory Delayed Recall | 20.43 (8.58) | 27.22 (3.80) | 3.79 | 0.0734 | 23.52 | 0.0005*** | 0.39 | 0.5474 |
| | Logical Memory Retention | 84.71 (25.65) | 93.65 (10.81) | 0.11 | 0.7510 | 5.14 | 0.0445* | 1.88 | 0.1975 |
| Visuospatial Memory | BVMT Immediate Recall | 23.71 (8.38) | 27.56 (5.32) | 2.13 | 0.168 | 5.16 | 0.0424* | 0.11 | 0.7487 |
| | BVMT Delayed Recall | 8.14 (4.22) | 11.11 (0.93) | 5.30 | 0.0385 * | 1.729 | 0.2130 | 0.05 | 0.8340 |
| | BVMT Retention | 0.89 (0.43) | 0.97 (0.04) | 2.04 | 0.1770 | 0.011 | 0.9190 | 0.75 | 0.4020 |
| | Visuospatial Working Memory | 7.00 (1.51) | 7.75 (1.04) | 2.42 | 0.144 | 1.53 | 0.2380 | 0.01 | 0.9380 |
| Associative Working Memory | Paired Associates Learning | 4.00 (0.76) | 4.62 (1.06) | 8.02 | 0.0142 * | 0.126 | 0.7280 | 0.14 | 0.710 |
| Spatial Short Term Memory Capacity | Spatial span | 5.62 (0.74) | 5.88 (1.36) | 0.70 | 0.4190 | 2.49 | 0.1390 | 0.18 | 0.6800 |
| Search Strategy | Self-ordered search | 7.25 (1.98) | 8.12 (1.55) | 6.15 | 0.0277 * | 6.54 | 0.0238* | 1.61 | 0.2263 |
| Attentional Processing | Feature Match | 110.25 (28.31) | 128.12 (27.66) | 1.57 | 0.2330 | 5.40 | 0.0370* | 0.29 | 0.601 |
